# The impact of genome-wide histocompatibility on liver transplantation outcomes

**DOI:** 10.1101/2024.07.16.24310481

**Authors:** M Semenova, V Liukkonen, S Markkinen, M Färkkilä, A Nordin, J Partanen, F Åberg, K Hyvärinen

**Author notes:** Corresponding author: Kati Hyvärinen, PhD, Associate professor, Finnish Red Cross Blood Service, Research and Development, Biomedicum Helsinki 1, Haartmaninkatu 8, 00290 Helsinki, FINLAND.

## Abstract

Liver transplantation (LT) is the standard treatment for end-stage liver diseases. However, the role of human leukocyte antigen (HLA)-matching in LT remains unclear. Immunological allograft injury and rejection are ongoing concerns, particularly when efforts are made to minimize immunosuppression. Although HLA matching currently has no established role in LT, interest in non-HLA compatibility in the field of transplantation is growing.

We compared 666 LT recipient-donor pairs and identified amino acid-changing genetic mismatches outside the HLA gene segment in different protein groups, and mismatches in 40 common gene deletions.

We evaluated the association between mismatches and LT outcomes by using adjusted Cox models for missense variant mismatches and deletion analyses. The primary endpoints were acute rejection, late rejection, graft loss, and overall survival. Statistical significance was set at a false discovery rate (FDR) <0.05.

Mismatches in missense variants coding for all proteins were associated with late rejection, with an adjusted hazard ratio (aHR) of 0.998 (95% confidence interval [CI]: 0.996–0.999; *P* = 0.011, FDR <0.05). Deletion mismatches tagged with rs11985201, rs2342606, rs2174926, and rs1944862 were identified as risk factors for LT outcomes. The sum of mismatches in deletion variants rs11985201, rs2342606, and rs1944862 was associated with time to acute rejection, with an aHR of 1.377 (95% CI: 1.139– 1.664, *P* = 0.001, FDR <0.05).

Genome-wide mismatches outside of the HLA region contribute to the risk of LT complications. Robust, large-scale studies are required to validate these results.

## 1. INTRODUCTION

Liver transplantation (LT) is the standard treatment for end-stage liver disease, acute liver failure, and some liver cancers. Survival after LT has improved in recent years, with more than 60% of recipients surviving for more than 10 years^1^. With modern immunosuppressive regimens, immunological complications, including acute rejection (AR) and chronic rejection, rarely lead to graft loss, although late AR is associated with inferior graft and recipient survival^2^. Recognition of the potential role of donor-specific antibodies (DSA)^3^, especially in minimizing immunosuppression, and reports of subclinical immunological damage and fibrosis in liver allografts^4^ have revived interest in immunology in recent years. However, most graft losses occur for non-immunological reasons^5^.

Cancer, cardiovascular events, renal failure, and infections are major causes of mortality after LT, and immunosuppressive medications predispose recipients to these events^5^. Thus, minimizing immunosuppression to improve survival is a major goal in post-LT management^6^. As the risk of complications varies among individuals, an individualized approach to immunosuppression management could improve post-LT outcomes.

The significance of human leukocyte antigens (HLA) in hematopoietic stem cell transplantation (HSCT)^7^ and kidney transplantation is well understood^8^. In the last decade, mismatches in genes located outside the HLA segment between recipients and donors have emerged as novel histocompatibility factors for HSCT^9,10^ and kidney transplantation^11–15^. For example, according to a recent study, kidney allograft survival is influenced by incompatibility with genes outside HLA^12^. In another recent study, researchers found that a genomic mismatch at the *LIMS1* locus is associated with the rejection of kidney allografts^11^.

The effect of HLA compatibility on LT outcomes is still debated, with studies showing mixed outcomes,^16–18^ and HLA matching not applied to organ allocation. There is a lack of data regarding the role of genome-wide compatibility in LT. Identifying novel risk factors for immunological and other important post-LT complications may facilitate risk stratification and individualized management^19^.

In this study, we analyzed the association between genome-wide incompatibility and time to AR, late AR, graft loss, and death in a single-center LT cohort of 666 recipient-donor pairs to identify novel genetic risk factors for LT outcomes.

## 2. STUDY SUBJECTS AND METHODS

### 2.1 Study cohort and design

The study conformed to the principles of the Declaration of Helsinki and was approved by the Ethics Committee of Helsinki University Hospital (HUS/155/2021), Finnish Medicines Agency Fimea (FIMEA/2021/004031), and the Abdominal Center, Helsinki University Hospital (HUS/155/2021/68). The study was conducted in accordance with the local legislation and institutional requirements. The ethics committee/institutional review board waived the requirement of written informed consent for participation from the participants or the participants’ legal guardians/next of kin because The Finnish Medicines Agency Fimea (previously the Finnish National Supervisory Authority for Welfare and Health) can authorize permission to study biological samples without informed consent if participants can not be reached and the study proposal is justifiable.

The characteristics of the recipients are presented in **Tables 1** and **2**. A flowchart of the study cohort is shown in **Supplementary Figure 1**. In total, 894 liver transplantation recipients who received a liver transplant between 2000 and 2018 and their respective first transplantation donors were included in the present study. DNA samples from both recipients and donors were extracted from whole blood samples at the Finnish Red Cross Blood Service, in Helsinki. After excluding individuals lacking DNA samples, the study cohort was genotyped into three cohorts: genotyping cohort I (286 donors), genotyping cohort II (30 recipients and 125 donors), and genotyping cohort III (1202 individuals). Individuals who failed to pass quality control (QC), had missing genotyping information, or were younger than 18 years were excluded.

**Table 1.**
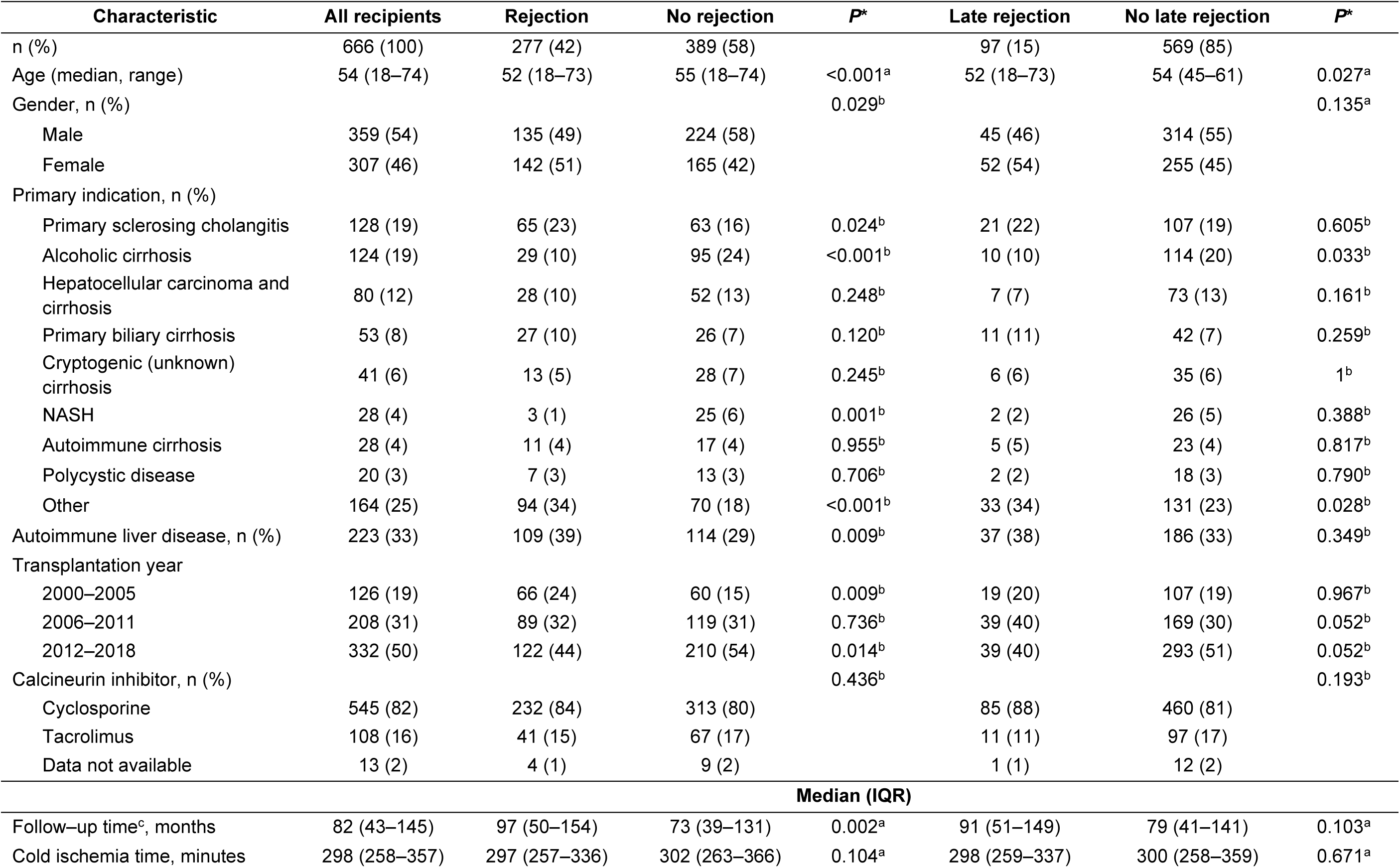
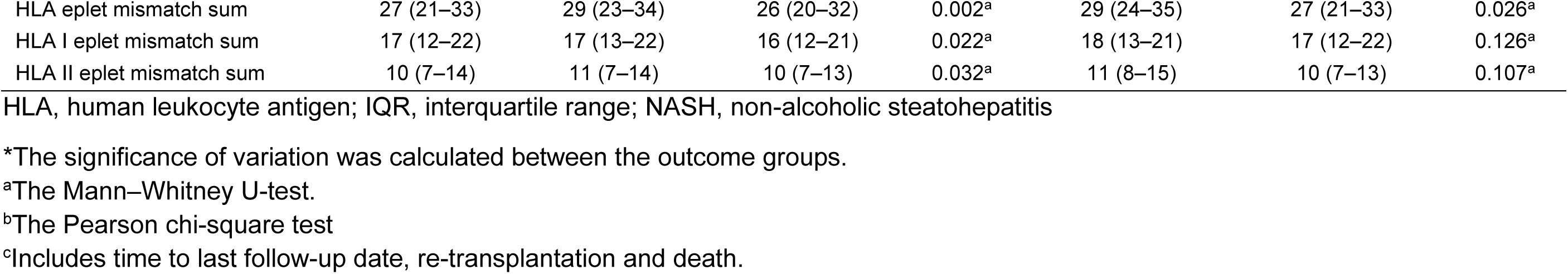
Characteristics of the study population for acute and late rejection groups.

| Characteristic | All recipients | Rejection | No rejection | *P* * | Late rejection | No late rejection | *P* * |
| --- | --- | --- | --- | --- | --- | --- | --- |
| n (%) | 666 (100) | 277 (42) | 389 (58) |  | 97 (15) | 569 (85) |  |
| Age (median, range) | 54 (18–74) | 52 (18–73) | 55 (18–74) | <0.001<sup>a</sup> | 52 (18–73) | 54 (45–61) | 0.027<sup>a</sup> |
| Gender, n (%) |  |  |  | 0.029<sup>b</sup> |  |  | 0.135<sup>a</sup> |
| Male | 359 (54) | 135 (49) | 224 (58) |  | 45 (46) | 314 (55) |  |
| Female | 307 (46) | 142 (51) | 165 (42) |  | 52 (54) | 255 (45) |  |
| Primary indication, n (%) |  |  |  |  |  |  |  |
| Primary sclerosing cholangitis | 128 (19) | 65 (23) | 63 (16) | 0.024<sup>b</sup> | 21 (22) | 107 (19) | 0.605<sup>b</sup> |
| Alcoholic cirrhosis | 124 (19) | 29 (10) | 95 (24) | <0.001<sup>b</sup> | 10 (10) | 114 (20) | 0.033<sup>b</sup> |
| Hepatocellular carcinoma and cirrhosis | 80 (12) | 28 (10) | 52 (13) | 0.248<sup>b</sup> | 7 (7) | 73 (13) | 0.161<sup>b</sup> |
| Primary biliary cirrhosis | 53 (8) | 27 (10) | 26 (7) | 0.120<sup>b</sup> | 11 (11) | 42 (7) | 0.259<sup>b</sup> |
| Cryptogenic (unknown) cirrhosis | 41 (6) | 13 (5) | 28 (7) | 0.245<sup>b</sup> | 6 (6) | 35 (6) | 1<sup>b</sup> |
| NASH | 28 (4) | 3 (1) | 25 (6) | 0.001<sup>b</sup> | 2 (2) | 26 (5) | 0.388<sup>b</sup> |
| Autoimmune cirrhosis | 28 (4) | 11 (4) | 17 (4) | 0.955<sup>b</sup> | 5 (5) | 23 (4) | 0.817<sup>b</sup> |
| Polycystic disease | 20 (3) | 7 (3) | 13 (3) | 0.706<sup>b</sup> | 2 (2) | 18 (3) | 0.790<sup>b</sup> |
| Other | 164 (25) | 94 (34) | 70 (18) | <0.001<sup>b</sup> | 33 (34) | 131 (23) | 0.028<sup>b</sup> |
| Autoimmune liver disease, n (%) | 223 (33) | 109 (39) | 114 (29) | 0.009<sup>b</sup> | 37 (38) | 186 (33) | 0.349<sup>b</sup> |
| Transplantation year |  |  |  |  |  |  |  |
| 2000–2005 | 126 (19) | 66 (24) | 60 (15) | 0.009<sup>b</sup> | 19 (20) | 107 (19) | 0.967<sup>b</sup> |
| 2006–2011 | 208 (31) | 89 (32) | 119 (31) | 0.736<sup>b</sup> | 39 (40) | 169 (30) | 0.052<sup>b</sup> |
| 2012–2018 | 332 (50) | 122 (44) | 210 (54) | 0.014<sup>b</sup> | 39 (40) | 293 (51) | 0.052<sup>b</sup> |
| Calcineurin inhibitor, n (%) |  |  |  | 0.436<sup>b</sup> |  |  | 0.193<sup>b</sup> |
| Cyclosporine | 545 (82) | 232 (84) | 313 (80) |  | 85 (88) | 460 (81) |  |
| Tacrolimus | 108 (16) | 41 (15) | 67 (17) |  | 11 (11) | 97 (17) |  |
| Data not available | 13 (2) | 4 (1) | 9 (2) |  | 1 (1) | 12 (2) |  |
| **Median (IQR)** |  |  |  |  |  |  |  |
| Follow-up time<sup>c</sup>, months | 82 (43–145) | 97 (50–154) | 73 (39–131) | 0.002<sup>a</sup> | 91 (51–149) | 79 (41–141) | 0.103<sup>a</sup> |
| Cold ischemia time, minutes | 298 (258–357) | 297 (257–336) | 302 (263–366) | 0.104<sup>a</sup> | 298 (259–337) | 300 (258–359) | 0.671<sup>a</sup> |
| HLA eplet mismatch sum | 27 (21–33) | 29 (23–34) | 26 (20–32) | 0.002<sup>a</sup> | 29 (24–35) | 27 (21–33) | 0.026<sup>a</sup> |
| HLA I eplet mismatch sum | 17 (12–22) | 17 (13–22) | 16 (12–21) | 0.022<sup>a</sup> | 18 (13–21) | 17 (12–22) | 0.126<sup>a</sup> |
| HLA II eplet mismatch sum | 10 (7–14) | 11 (7–14) | 10 (7–13) | 0.032<sup>a</sup> | 11 (8–15) | 10 (7–13) | 0.107<sup>a</sup> |

HLA, human leukocyte antigen; IQR, interquartile range; NASH, non-alcoholic steatohepatitis
\*The significance of variation was calculated between the outcome groups.
<sup>a</sup>The Mann–Whitney U-test.
<sup>b</sup>The Pearson chi-square test
<sup>c</sup>Includes time to last follow-up date, re-transplantation and death.

**Table 2.** Characteristics of the study population for graft loss and overall survival groups.

| Characteristic | All recipients | Graft loss | No graft loss | *P*<sup>*</sup> | Deceased | Alive | *P*<sup>*</sup> |
| --- | --- | --- | --- | --- | --- | --- | --- |
| n (%) | 666 (100) | 42(6) | 624 (94) |  | 163 (24) | 503 (76) |  |
| Age (median, range) | 54 (18–74) | 50 (18–64) | 54 (18–74) | 0.007<sup>a</sup> | 58 (18–74) | 52 (18–73) | <0.001<sup>a</sup> |
| Gender, n (%) |  |  |  | 0.494<sup>b</sup> |  |  | 0.543<sup>b</sup> |
| Male | 359 (54) | 20 (48) | 339 (54) |  | 84 (52) | 275 (55) |  |
| Female | 307 (46) | 22 (52) | 285 (46) |  | 79 (48) | 228 (45) |  |
| Primary indication, n (%) |  |  |  |  |  |  |  |
| Primary sclerosing cholangitis | 128 (19) | 12 (29) | 116 (19) | 0.166<sup>b</sup> | 23 (14) | 105 (21) | 0.073<sup>b</sup> |
| Alcoholic cirrhosis | 124 (19) | 3 (7) | 121 (19) | 0.077<sup>b</sup> | 26 (16) | 98 (19) | 0.373<sup>b</sup> |
| Hepatocellular carcinoma and cirrhosis | 80 (12) | 2 (5) | 78 (13) | 0.212<sup>b</sup> | 25 (15) | 55 (11) | 0.173<sup>b</sup> |
| Primary biliary cirrhosis | 53 (8) | 5 (12) | 48 (8) | 0.495<sup>b</sup> | 11 (7) | 42 (8) | 0.624<sup>b</sup> |
| Cryptogenic (unknown) cirrhosis | 41 (6) | 3 (7) | 38 (6) | 1<sup>b</sup> | 16 (10) | 25 (5) | 0.040<sup>b</sup> |
| NASH | 28 (4) | 0 (0) | 28 (4) | 0.315<sup>b</sup> | 7 (4) | 21 (4) | 1<sup>b</sup> |
| Autoimmune cirrhosis | 28 (4) | 1 (2) | 27 (4) | 0.833<sup>b</sup> | 7 (4) | 21 (4) | 1<sup>b</sup> |
| Polycystic disease | 20 (3) | 0 (0) | 20 (3) | 0.477<sup>b</sup> | 2 (1) | 18 (4) | 0.206<sup>b</sup> |
| Other | 164 (25) | 16 (38) | 148 (24) | 0.056<sup>b</sup> | 46 (28) | 118 (23) | 0.262<sup>b</sup> |
| Autoimmune liver disease, n (%) | 223 (33) | 20 (48) | 203 (33) | 0.066<sup>b</sup> | 48 (29) | 175 (35) | 0.246<sup>b</sup> |
| Transplantation year |  |  |  |  |  |  |  |
| 2000–2005 | 126 (19) | 12 (29) | 114 (18) | 0.148<sup>b</sup> | 51 (32) | 75 (15) | <0.001<sup>b</sup> |
| 2006–2011 | 208 (31) | 15 (36) | 193 (31) | 0.634<sup>b</sup> | 58 (36) | 150 (30) | 0.200<sup>b</sup> |
| 2012–2018 | 332 (50) | 15 (36) | 317 (51) | 0.083<sup>b</sup> | 54 (33) | 278 (55) | <0.001<sup>b</sup> |
| Calcineurin inhibitor, n (%) |  |  |  | 0.071<sup>b</sup> |  |  | 0.393<sup>b</sup> |
| Cyclosporine | 545 (82) | 38 (90) | 507 (81) |  | 135 (83) | 410 (82) |  |
| Tacrolimus | 108 (16) | 2 (5) | 106 (17) |  | 22 (13) | 86 (17) |  |
| Data not available | 13 (2) | 2 (5) | 11 (2) |  | 6 (4) | 7 (1) |  |
| **Median (IQR)** |  |  |  |  |  |  |  |
| Follow-up time<sup>c</sup>, months | 82 (43–145) | 34(4–77) | 88 (47–147) | <0.001<sup>a</sup> | 39 (13–103) | 93 (55–150) | <0.001<sup>a</sup> |
| Cold ischemia time, minutes | 298 (258–357) | 284 (253–342) | 299 (258–359) | 0.573<sup>a</sup> | 316 (267–375) | 294 (257–345) | 0.013<sup>a</sup> |
| HLA eplet mismatch sum | 27 (21–33) | 27 (21–32) | 27 (21–33) | 0.857<sup>a</sup> | 27 (21–35) | 27 (21–33) | 0.812<sup>a</sup> |
| HLA I eplet mismatch sum | 17 (12–22) | 17 (11–21) | 17 (12–22) | 0.443<sup>a</sup> | 17 (12–23) | 17 (12–21) | 0.588<sup>a</sup> |
| HLA II eplet mismatch sum | 10 (7–14) | 10 (7–14) | 10 (7–14) | 0.649<sup>a</sup> | 10 (7–14) | 10 (7–14) | 0.895<sup>a</sup> |

HLA, human leukocyte antigen; IQR, interquartile range; NASH, non-alcoholic steatohepatitis
\*The significance of variation was calculated between the outcome groups.
<sup>a</sup>The Mann–Whitney U-test.
<sup>b</sup>The Pearson chi-square test
<sup>c</sup>Includes time to last follow-up date, re-transplantation and death.

All ARs were confirmed by biopsy using the BANFF scheme^20^. Late rejection was defined as AR diagnosed more than 90 days after transplantation. Primary or secondary LT indications for primary sclerosing cholangitis, primary biliary cholangitis, or autoimmune hepatitis were categorized as autoimmune liver disease.

### 2.2 Genotyping and genotype imputation

The flow of the genetic variants in the pre- and post-imputation processes is shown in **Supplementary Figure 2**. Genotyping was performed at the Finnish Institute of Molecular Medicine (Helsinki, Finland). Cohort I was genotyped using the Illumina Infinium™ Global Screening Array (GSA)-24 v3.0 BeadChip + Multi-Disease (MD) drop-in customized with 34,191 Finnish variants. Cohorts II and III were genotyped using Illumina Infinium™ GSA-24 v2.0 + MD bead chip. Prior to the genotype imputation, results in build GRCh37 were lifted over to build GRCh38^21^. Genotype imputation was performed with Beagle v4.1 program using the THL Biobank’s SISu v3 reference panel according to Palta *et al.*^22^ (https://www.protocols.io/view/genotype-imputation-workflow-v3-0-xbgfijw?step=4&comment_id=86624.) In post-imputation QC, individuals with missing genotype >5%, variants with missing data rate >10%, minor allele frequency <1%, and imputation INFO score <0.6 were excluded. A total of 8,706,949 common variants were available for the analysis.

### 2.3 HLA imputation and eplet mismatches

Imputation of alleles of HLA-A, -B, -C, -DRB1, -DQA1, -DQB1, and -DPB1 was performed according to Ritari *et al.*^23^ using the R package HIBAG in RStudio version 3.6.3. (https://github.com/FRCBS/HLA-imputation). The use of the Finnish HLA imputation reference panel has been shown to result in the accurate imputation of HLA alleles.

HLA eplet mismatches between recipients and donors were calculated for the imputed HLA-A, -B, -C, -DRB, -DQB, and -DPB alleles using HLAMatchMaker (http://www.epitopes.net/)^24^. The HLA-A, -B, and -C alleles were matched using the <u>ABC Eplet Matching Program V4.0</u>, and the HLA-DRB, -DPB, -DQA, and -DQB alleles were matched using the <u>DRDQDP Eplet Matching Program V3.1</u>. These programs were downloaded from http://www.epitopes.net/downloads.html (downloaded June 23, 2022).

### 2.4 Missense variant mismatch analyses

Prior to mismatch analyses, variants coding for the major histocompatibility complex region on chromosome 6 and both sex chromosomes were excluded, leaving only autosomal non-HLA variants in the dataset. **Supplementary Figure 3** shows the flow of the variants. Functional annotation of variants was performed using the Ensemble Variant Effect Predictor^25^ release 106 (https://www.ensembl.org/info/docs/tools/vep/script/index.html). The transcripts of all transmembrane and secreted proteins, transmembrane proteins, and liver-related proteins were retrieved from UniProt 84 (https://www.uniprot.org/). The filtering operator “consequence is missense_variants” was used (“all proteins”). The term missense variant refers to a nucleotide polymorphism that changes the encoded amino acid in a protein.

The query strings for the different protein groups were as follows:

1. Transmembrane and secreted proteins (annotation:(type:transmem) OR locations:(location:“Secreted [SL-0243]”) OR keyword:“Transmembrane [KW-0812]”) AND reviewed:yes AND organism:“Homo sapiens (Human) [9606]“
2. Transmembrane proteins (annotation:(type:transmem) OR keyword:“Transmembrane [KW-0812]”) AND reviewed:yes AND organism:“Homo sapiens (Human) [9606]“
3. Liver-related proteins annotation:(type:“tissue specificity” liver) AND reviewed:yes AND organism:“Homo sapiens (Human) [9606]“

The directionality of the mismatch is from donor to recipient; consequently, a new allele is introduced from donor to recipient. We compared the recipient and donor genomes pairwise, and calculated the missense variant mismatch sums between the genomes of four different protein groups: missense variants coding for 1) all proteins, 2) transmembrane and secreted proteins, 3) transmembrane proteins only, and 4) liver-related proteins.

Cox proportional hazards models were used to investigate the effects of missense variant mismatches on the time to AR, late rejection, graft loss, and death. We analyzed the missense variant mismatch sums and quartiles of missense variant mismatch sums in each protein group. All models were adjusted for recipient and donor sex, recipient and donor age, cold ischemia time, HLA I eplet mismatch, HLA II eplet mismatch, year of transplantation, autoimmune liver disease, and initial calcineurin inhibitor type. Censoring was performed at the end of the follow-up period for retransplantation or death. Kaplan–Meier plots were used to visualize the AR-, late rejection-, graft loss-free, and overall survival of the recipients.

### 2.5 Deletion analyses

The study protocol described by Markkinen *et al.*^26^ was followed. Forty genetic variant tagging (*i.e*., variants known to be in strong linkage disequilibrium) gene deletions and their frequencies in the study cohort are shown in **Supplementary Table S1**. Deletion mismatch was defined as a situation in which the recipient was homozygous for a deletion-tagging allele and received a liver from a donor who was either homozygous or heterozygous for the other allele.

We analyzed the effects of homozygous mismatches in the 40 deletion-tagged variants by using adjusted Cox proportional hazards models. Censoring was performed at the end of the follow-up period for retransplantation or death. The dependent variables were the time to AR, late AR, graft loss, and death. We investigated deletion-tagging variant mismatches separately and the quartiles of the sum of deletion-tagging variant mismatches. In addition, we analyzed the sums of mismatches in rs11985201, rs2342606, rs2174926, and rs1944862 (variants with nominal P<0.05, with n values of homozygous deletion-tagging variant mismatches >10). All models were adjusted for donor and recipient age, donor and recipient sex, cold ischemia time, HLA I eplet mismatch, HLA II eplet mismatch, year of transplantation, autoimmune liver disease, and initial calcineurin inhibitor type. Kaplan–Meier survival curves were used to visualize AR-, late rejection-, graft loss-free, and overall survival of the recipients. Full Cox models and Kaplan–Meier plots are presented for variants with nominal *P* <0.05, with n values of homozygous deletion-tagging variant mismatches >10 (**Supplementary Table S1**).

### 2.6 Expression quantitative trait loci and regulatory element annotations

Expression quantitative trait loci (eQTL) results, that is, associations of these genetic variants with the gene expression levels of other proteins, were searched for four deletion-tagging variants: rs11985201, rs2342606, rs2174926, and rs1944862, using FIVEx^27^ https://fivex.sph.umich.edu/ and eQTLGen phase I^28^ https://www.eqtlgen.org/phase1.html. Regulatory elements were identified using the RegulomeDB^29^ database (https://regulomedb.org/regulome-search/. The Regulome DB rank varies from 1(a-f) to 7, and the score ranges from 0 to 1 (with 1 being the most reliable result). Regulatory motifs and annotations were explored using Haploreg^30^ v4.2 tool https://pubs.broadinstitute.org/mammals/haploreg/haploreg.php.

### 2.7 Other statistical analyses

The normality of the variable distributions was evaluated using the Shapiro–Wilk normality test. In **Tables 1** and **2**, the variation between the outcome groups was analyzed using Pearson’s chi-square test for categorical data (recipient sex, primary indication, autoimmune liver disease, and initial calcineurin inhibitor type) and the nonparametric Mann-Whitney U test for non-normally distributed data (recipient age, follow-up time, cold ischemia time, HLA eplet mismatch sum, HLA I eplet mismatch sum, and HLA II eplet mismatch sum). Statistical significance was set at *P* <0.05.

To counteract the multiple comparison problem in mismatch analyses, the false discovery rate (FDR) was determined and FRD <0.05, which was considered statistically significant.

All analyses were performed using R v3.6.3^31^.

The study follows the relative reporting STROBE-guidelines.

## 3. RESULTS

### 3.1 Characteristics of the study cohort

The characteristics of the recipients in the study cohort are presented in **Tables 1** and **2**. During the follow-up period (median interquartile range [IQR], 7 [4–12] years), 42% of the 666 recipients experienced AR, 15% experienced late rejection, 6% experienced graft loss, and 24% died. Women experienced AR more often than men did (51% vs. 49%, *P* = 0.029). The median (IQR) time to AR was 21 (11–87) days. The median (IQR) time to late rejection was 11 (5–23) months. The median (IQR) time to graft loss was 34 (4–77) months and the median (IQR) time to death was 3 (1–9) years.

**Figure 1.**
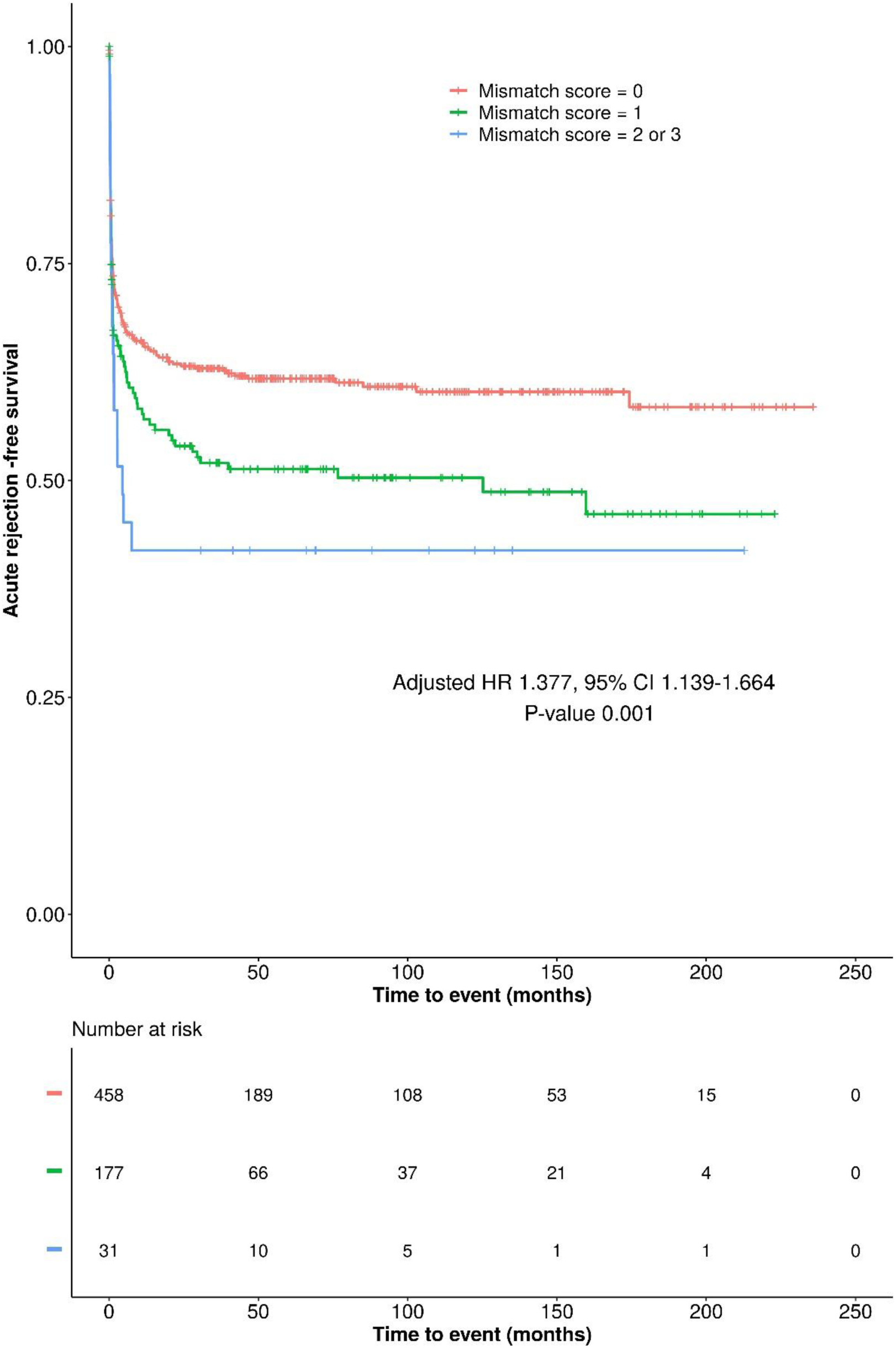
Kaplan–Meier plot displaying the effect of sum of mismatches in deletion-tagging variants rs11985201, rs2342606, and rs1944862 on acute rejection-free survival. Survival curves represent the mismatch score groups 0,1, and combined scores 2 and 3. Under the survival curve, a risk table is displayed with the number of recipients in each group that are at risk of acute rejection. The presented hazard ratios, confidence intervals, and P values were calculated using the Cox proportional hazards model. The model was adjusted for recipient and donor age, recipient and donor sex, cold ischemia time, HLA I eplet mismatch, HLA II eplet mismatch, year of transplantation, autoimmune liver disease, and initial calcineurin inhibitor type. CI, confidence interval; HR, hazard ratio.

### 3.2 Missense variant mismatch analyses

After excluding sex chromosomes, major histocompatibility complex regions, and non-missense variants, 28,225 missense variants were available for mismatch analysis. Of these missense variants, 9,337 coded for both transmembrane and secreted proteins, 6,963 for transmembrane proteins only, and 3,199 for liver-related proteins (**Supplementary Figure S3**). **Supplementary Figures S4–S7** show the variance in missense variants in the LT outcome groups. The results of the Cox proportional hazards models for both the sum and quartiles of missense variant mismatches are presented in **Table 3**. **Supplementary Figure S8** shows the effects of quartiles of missense variant mismatches of all proteins on late rejection-free survival. **Supplementary Figure S9** shows the effects of quartiles of missense variant mismatches in the transmembrane proteins on graft loss-free survival. All models were adjusted for recipient and donor age, recipient and donor sex, cold ischemia time, HLA I eplet mismatch, HLA II eplet mismatch, year of transplantation, autoimmune liver disease, and initial calcineurin inhibitor type. We observed a negative association between both the sum and quartiles of mismatches in missense variants in all proteins and late rejection time (HR 0.998, 95% CI 0.996–0.999, *P* = 0.011, FDR <0.05; HR 0.812, 95% CI 0.674–0.977, *P* = 0.028, FDR >0.05, respectively).

**Table 3.** Summary statistics of adjusted Cox proportional hazards models for four different missense variant groups.

| Endpoint | Missense variants | Cox proportional hazards model |  |  |  |
| --- | --- | --- | --- | --- | --- |
|  |  | Sum of mismatches |  | Quartiles of mismatches |  |
|  |  | HR (95% CI) | P value | HR (95% CI) | P value |
| Time to acute rejection | All proteins | 1.000(0.999–1.001) | 0.891 | 1.003(0.900–1.118) | 0.956 |
|  | Transmembrane and secreted proteins | 1.000(0.998–1.002) | 0.820 | 1.041(0.934–1.161) | 0.466 |
|  | Transmembrane proteins | 1.000(0.998–1.002) | 0.787 | 1.021(0.916–1.138) | 0.708 |
|  | Liver-related proteins | 0.999(0.995–1.003) | 0.484 | 0.985(0.883–1.099) | 0.791 |
| Time to late rejection | All proteins | 0.998(0.996–0.999) | 0.011* | 0.812(0.674–0.977) | 0.028 |
|  | Transmembrane and secreted proteins | 0.997(0.994–1.000) | 0.070 | 0.890(0.740–1.071) | 0.218 |
|  | Transmembrane proteins | 0.997(0.993–1.001) | 0.130 | 0.859(0.714–1.034) | 0.109 |
|  | Liver-related proteins | 0.994(0.987–1.001) | 0.122 | 0.937(0.779–1.127) | 0.489 |
| Time to graft loss | All proteins | 0.999(0.997–1.002) | 0.657 | 0.866(0.649–1.156) | 0.329 |
|  | Transmembrane and secreted proteins | 0.996(0.991–1.001) | 0.164 | 0.747(0.559–1.000) | 0.050 |
|  | Transmembrane proteins | 0.995(0.989–1.001) | 0.125 | 0.740(0.551–0.994) | 0.046 |
|  | Liver-related proteins | 1.006(0.995–1.016) | 0.297 | 1.163(0.869–1.556) | 0.309 |
| Overall survival | All proteins | 0.999(0.998–1.001) | 0.226 | 0.907(0.784–1.049) | 0.189 |
|  | Transmembrane and secreted proteins | 0.999(0.996–1.001) | 0.358 | 0.897(0.777–1.036) | 0.138 |
|  | Transmembrane proteins | 0.999(0.996–1.002) | 0.647 | 0.934(0.809–1.077) | 0.348 |
|  | Liver-related proteins | 0.997(0.991–1.002) | 0.271 | 0.956(0.828–1.105) | 0.546 |

MHC region and sex chromosomes excluded. CI, confidence interval; HR, hazard ratio. Models were adjusted with donor and recipient age, donor and recipient sex, cold ischemia time, HLA I eplet mismatch, HLA II eplet mismatch, year of transplantation, autoimmune liver disease and initial calcineurin inhibitor type.
\* False detection rate <0.05.

### 3.3 Deletion analyses

The results of the adjusted Cox proportional hazards models for the 40 deletion-tagging variants are summarized separately in **Supplementary Tables S2–S5** for AR, late rejection, graft loss times, and overall survival, respectively.

The results from the adjusted Cox proportional hazards models showed that a deletion mismatch in variants rs11985201 (tagging deletion of *ADAM3A/ADAM5P*) and rs2342606 (tagging deletion of *LOC642521* and *LOC642538*) was associated with AR time, with an HR of 1.483 (95% CI: 1.066–2.062, *P* = 0.019, **Supplementary Figure S10**, **Supplementary Table S6**) and 1.373 (95% CI: 1.011–1.865, *P* = 0.042; **Supplementary Figure S11**, **Supplementary Table S7**), respectively. In addition, late rejection time was associated with a deletion mismatch in rs1944862 (tagging deletion of *OR4P1P*) with an HR of 2.341 (95% CI: 1.326–4.130, *P* = 0.003; **Supplementary Figure S12**, **Supplementary Table S8**), and graft loss time with a deletion mismatch in rs2174926 (tagging deletion of *LOC442434*) with an HR of 2.332 (95% CI: 1.145–4.752, *P* = 0.020, **Supplementary Figure S13**, **Supplementary Table S9**). However, none of the *P* values reached statistical significance (FDR <0.05) (**Supplementary Tables S2–S5**).

**Supplementary Figures S14–S17** show the effect of quartiles of the overall deletion mismatch sums on the LT endpoints. No association was found between the quartiles of the overall deletion mismatch sums and LT endpoints.

Next, we analyzed the risk sums of deletion-tagging variants with nominal P-values of <0.05. The results of the adjusted Cox proportional hazards models for the sums of variant mismatches are presented in **Table 4**. The sum of mismatches in rs11985201 and rs2342606 was associated with AR time with an HR of 1.414 (95% CI: 1.132– 1.767, *P* = 0.002, FDR >0.05). The sum of mismatches in rs11985201, rs2342606, and rs1944862 was associated with an increased risk of AR, with an HR of 1.377 (95% CI: 1.139–1.664, *P* = 0.001, FDR <0.05), and an increased risk of LR, with an HR of 1.512 (95% CI: 1.124–2.033, *P* = 0.006, FDR >0.05). In addition, the sum of mismatches in rs11985201, rs2342606, rs1944862, and rs2174926 was associated with AR time with an HR of 1.261 (95% CI: 1.069–1.489, *P* = 0.006, FDR >0.05) and an increased risk of LR, with an HR of 1.442 (95% CI: 1.104–1.884, *P* = 0.007, FDR >0.05). The effect of sum of mismatches in deletion-tagging variants rs11985201, rs2342606, and rs1944862 on acute rejection-free survival is displayed in **Figure 1**.

**Table 4.** Adjusted Cox proportional hazards models for sums of mismatches in risk variants across four endpoints.

| Independent variable (risk type) | AR |  | LR |  | GL |  | OS |  |
| --- | --- | --- | --- | --- | --- | --- | --- | --- |
|  | HR (95% CI) | P value | HR (95% CI) | P value | HR (95% CI) | P value | HR (95% CI) | P value |
| rs11985201 (AR risk) | 1.483(1.066–2.062) | 0.019 | 1.196(0.676–2.114) | 0.538 | 1.621(0.739–3.558) | 0.228 | 0.741(0.440–1.247) | 0.259 |
| rs2342606 (AR risk) | 1.373(1.011–1.865) | 0.042 | 1.530(0.926–2.528) | 0.097 | 0.290(0.069–1.212) | 0.090 | 0.724(0.448–1.170) | 0.187 |
| rs1944862 (LR risk) | 1.355(0.897–2.048) | 0.149 | 2.341(1.326–4.130) | 0.003 | 0.686(0.160–2.946) | 0.613 | 1.010(0.542–1.883) | 0.975 |
| rs2174926 (GL risk) | 0.989(0.709–1.380) | 0.948 | 1.173(0.681–2.020) | 0.566 | 2.332(1.145–4.752) | 0.020 | 1.329(0.887–1.990) | 0.168 |
| rs11985201, rs2342606 (AR risk variant sum) | 1.414(1.132–1.767) | 0.002 | 1.356(0.936–1.963) | 0.107 | 0.852(0.431–1.682) | 0.644 | 0.735(0.518–1.044) | 0.085 |
| rs11985201, rs2342606, and rs1944862 (AR+LR risk variant sum) | 1.377(1.139–1.664) | 0.001 | 1.512(1.124–2.033) | 0.006 | 0.829(0.457–1.503) | 0.536 | 0.789(0.581–1.071) | 0.128 |
| rs11985201, rs2342606, rs1944862, and rs2174926 (AR+LR+GL risk variant sum) | 1.261(1.069–1.489) | 0.006 | 1.442(1.104–1.884) | 0.007 | 1.186(0.755–1.862) | 0.458 | 0.934(0.733–1.192) | 0.585 |

AR, acute rejection; CI, confidence interval; GL, graft loss; HR, hazard ratio; LR, late rejection; OS, overall survival.

### 3.4 Expression quantitative trait loci analyses and regulatory element annotation

Genetic variants that tag gene deletions may have additional functions, such as regulating the expression levels of other genes, possibly affecting LT outcomes. Therefore, we screened public databases to obtain information on the regulatory functions of the deletion-tagging variants rs11985201, rs2342606, rs2174926, and rs1944862.

The *cis*-eQTL results for the deletion-tagged rs11985201, rs2342606, rs1944862, and rs2174926 variants in the FIVEx and eQTLGen phase I databases are shown in **Table 5**. Using the FIVEx browser, we found that rs2342606 was associated with increased expression of TMEM254 antisense RNA 1 (*TMEM252-AS1*) in several different tissues. In the eQTLGen phase I database, variant rs2342606 was associated with altered expression of the following genes: *TMEM254*, *TMEM254-AS1*, *FAM22B*, *RP11-119F19.2*, *FAM213A*, *SFTPD*, *RP11-137H2.4,* and *RP11-506M13.3*.

**Table 5.**
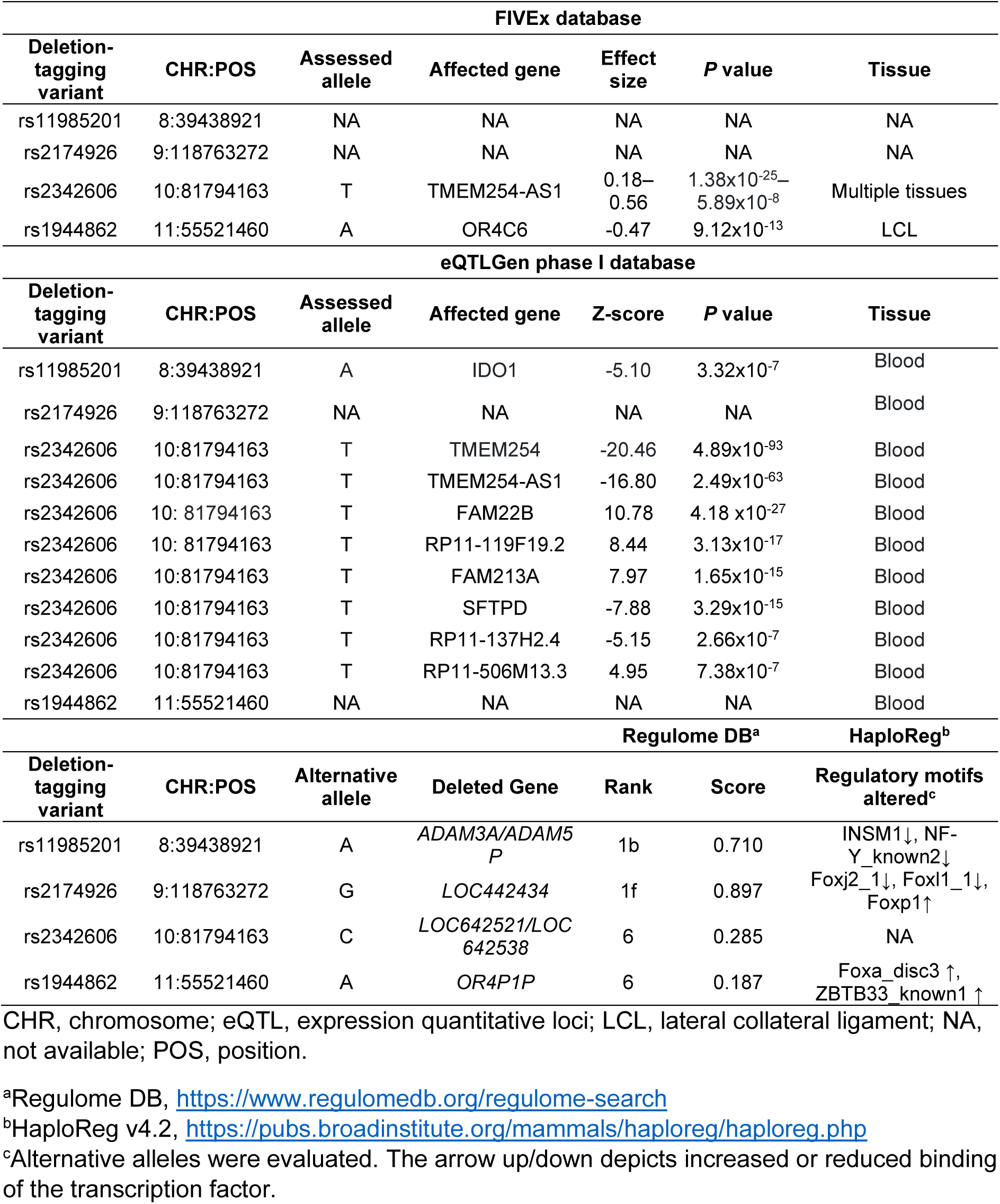
*cis-*eQTL results of deletion-tagging variants rs11985201, rs2342606, rs2174926, and rs1944862.

In the FIVEx database, rs11985201 was associated with decreased expression of the olfactory receptor family 4 subunit C member 6 (*OR4C6*) protein-coding gene in the lateral collateral ligament tissue. Using the eQTLGen phase I browser, we found that the variant rs11985201 was associated with decreased expression levels of the indoleamine 2,3-dioxygenase 1 (*IDO1*) gene in blood cells.

Regulome DB and HaploReg tools were used to identify regulatory elements (**Table 5**). Two variants, rs11985201 and rs2174926, showed high RegulomeDB rank and score, both of which are prediction scores for the quality of supporting evidence in the literature on the regulatory role of variants. The deletion-tagging variant rs11985201 is associated with reduced binding of the zinc-finger transcription factor (INSM1) and nuclear transcription factor Y (NF-Y_known2). The deletion-tagging variant rs2174926 decreased the binding of forkhead box J2 (Foxj2_1) and forkhead box L1 (Foxl1_1) and increased the binding of forkhead box P1 (Foxp1) transcription factors. The deletion-tagging variant rs1944862 was associated with increased binding of forkhead box A (Foxa_disc3), zinc finger, and btb domain-containing 33 (ZBTB33_known1) transcription factors.

## 4. DISCUSSION

In the present study, we investigated the association between LT endpoints and genome-wide mismatches in 666 LT recipient–donor pairs. We analyzed the mismatches in missense variants changing the amino acid sequence of the encoded protein and, additionally, in 40 common deletion-tagging variants. The sum of mismatches in rs11985201, rs2342606, and rs1944862 deletion-tagging variants was associated with an increased risk of AR (FDR <0.05) and could be a novel histocompatibility factor in LT.

Genetic differences between recipients and donors have been identified as important factors in allograft rejection and have been a topic of active research over the last decade^11–13^. HLA allele matching is well established in kidney and hematopoietic stem cell transplantation^32^. However, the importance of HLA matching between LT recipients and donors remains controversial^18,33–37^. Navarro *et al.* analyzed 29 675 first-time transplants and found no clinically significant effect of HLA matching on graft survival^18^. In contrast, a recent large retrospective study by Nabulsi *et* al. found that HLA mismatch is associated with increased acute rejection episodes and reduced graft survival^17^. A recent study found that HLA-A mismatch was associated with an increased risk of allograft loss after LT^34^, whereas a meta-analysis found a significant association between HLA-C mismatch and an increased risk of AR in LT^37^. In the present study, HLA class I eplet matching in certain deletion-tagged variants was associated with a higher risk of AR. However, the result was not statistically significant (FDR <0.05). Forner *et al.* reported no statistically significant association between HLA eplet mismatches and the risk of AR^38^.

In addition to HLA mismatches, genome-wide incompatibility between recipients and donors has emerged as a potential risk factor for kidney transplantation^11–14^. A study conducted by Reindl-Schwaighofer *et al.* demonstrated the effects of genome-wide genetic incompatibility between recipients and donors on graft survival during kidney transplantation^12^. The results indicated that mismatches in transmembrane and secreted proteins were associated with an increased risk of graft loss, independent of HLA^12^. Markkinen *et al.* in 2022 reported that an increasing number of mismatches in kidney-related proteins increases the risk of AR after kidney transplantation^26^. Studies investigating the importance of non-HLA genetic factors in LT outcomes are limited.

In this study, we did not observe a statistically significant association between missense variant mismatches and LT outcomes, except for late rejection time and missense variant mismatches in all protein group. Contrary to reports of kidney transplantation studies, the increased number of mismatches functioned as a protective factor against late rejections. However, the effect size was negligible, suggesting a limited practical application and the need for confirmation in a larger study.

Mismatches in common gene deletions introduce foreign protein structures into the recipients, which may cause an alloimmune reaction. The production of DSAs may mitigate negative post-LT outcomes. A recent study by Steers *et al.* (2019) found an association between kidney allograft rejection and genomic mismatch in *LIMS1* deletion^11^. These results were not confirmed in a Finnish patient cohort study by Markkinen *et al.*, who found an association between AR and mismatches in deletions at the complement factor H-related protein (CFHR) locus^26^. This study did not confirm any association between LIMS and CFHR deletion. However, we found associations between the four other deletion-tagging variants and the outcomes of LT. Specifically, we observed a significant association between the sum of mismatches in the rejection risk variants and AR. These results, together with those of HSCT^9^, suggest that deletion mismatches, in general, predispose patients to graft incompatibility, but that predominant deletions vary between transplant types and populations. The liver has a tolerogenic microenvironment that enables the regulation of local immune responses^39^. This may result in a higher tolerance to immunogenetic incompatibility in LT than in the other transplantation types. Further studies are required to determine the role of deletion mismatches and how they lead to a higher risk of rejection.

The lack of certain important immune regulatory proteins, such as CFHR1, may have a negative effect on transplantation outcomes. Furthermore, genetic variants may have various regulatory functions that affect LT outcomes. They can, *e.g*., regulate the expression levels of genes and affect the binding motifs of transcription factors. The variant rs11985201 downregulated the expression of IDO1, an important immunomodulatory enzyme, and reduced the binding of INSM1 and NF-Y_known2 transcription factors. rs2342606 tags the deletion of *LOC642521/LOC642538* but does not affect any known binding motifs. The deletion-tagging variant rs2174926 is associated with altered binding of several FOX transcription factors. The rs11985201 and rs1944862 variants were included in the top 15,000 methylation quantitative train loci identified in the human liver^40^, indicating that they were associated with DNA methylation.

This study has several strengths and limitations. One of the strengths of this study was the reasonable cohort size collected from a single center, which reduced heterogeneity. The LTs in the present study were from a relatively long period (19 years). We compensated for this potential effect by selecting transplantation year as one of the covariates. The cohort, albeit one of the largest published thus far for genome analyses, included only 666 recipient-donor pairs, which enabled the recognition of only relatively large effect sizes for the P-values to pass the correction for multiple comparisons. Larger collaborative studies are required to overcome this issue. However, to the best of our knowledge, this is the first large-scale single-center study to investigate the impact of genome-wide compatibility on the time to AR, late rejection, graft loss, and death in LT. A single-center study in a relatively homogeneous Finnish population has clear advantages by limiting the heterogeneity in patient demographics, treatments, and genetic background.

In conclusion, genetic incompatibility between recipients and donors at the whole-genome level may be a significant histocompatibility factor that affects LT outcomes. Robust large-scale studies are required to confirm the role of genome-wide mismatches and common variants in liver transplantation.

## Abbreviations

aHR: adjusted hazard ratio
AR: Acute rejection
CI: Confidence interval
CHR: Chromosome
DNA: Deoxyribonucleic acid
DSA: Donor-specific antibody
eQTL: Expression quantitative trait loci
FDR: False discovery rate
GSA: Global screening array
HLA: Human leukocyte antigen
HSCT: hematopoietic stem cell transplantation
IQR: Interquartile range
LCL: Lateral collateral ligament
LT: Liver transplantation
MAF: Minor allele frequency
MD: Multi-Disease
NA: Not available
POS: Position
QC: Quality control

## ACKNOWLEDGEMENTS

The reference data for genotype imputation was obtained from the THL Biobank (study number: BB2019_12). We thank all study participants for their generous participation in the biobank research.

## DATA AVAILABILITY STATEMENT

Individual genotype data are not publicly available because of restrictions issued by the ethical committee and current legislation in Finland, which do not allow the distribution of pseudonymized personal data, including genetic and clinical data.

Analysis scripts are available at https://github.com/FRCBS/Liver_tx_gw_mismatch_public.

## SUPPORTING INFORMATION STATEMENT

Additional supporting information may be found online in the Supporting Information section.

## AUTHOR CONTRIBUTION

MS, SM, JP, and KH designed this study. MS and KH performed the genome data analysis. VL, FÅ, AN, and MF provided clinical data and clinical expertise. MS, SM, JP, FÅ, and KH interpreted the data. MS and VL drafted the manuscript. All authors have read, accepted, and contributed to the final version of the manuscript.

