## Supplementary data for "The impact of genome-wide histocompatibility on liver transplantation outcomes"

Short title: Genome-wide mismatches in liver transplantation

Corresponding author:

Kati Hyvärinen, PhD, Associate professor

Finnish Red Cross Blood Service

Research and Development

Biomedicum Helsinki 1, Haartmaninkatu 8, 00290 Helsinki, FINLAND

**Supplementary Table S1. Number of deletion-tagging variant mismatches in the study cohort.**

| CHR | Deletion-tagging variant | A1 | A2 | MAF | Number of mismatches[a] (%) | | | | | | | | Affected genes[b] |
|---|---|---|---|---|---|---|---|---|---|---|---|---|---|
| | | | | | No rejection (n = 389) | Rejection (n = 277) | No graft loss (n = 624) | Graft loss (n = 42) | Alive (n = 503) | Deceased (n = 163) | No late rejection (n = 569) | Late rejection (n = 97) | |
| 1 | rs10927864 | G | C | 0.27 | 28 (7) | 16 (6) | 42 (7) | 2 (5) | 33 (7) | 11 (7) | 38 (7) | 6 (6) | RPS16P1 |
| 1 | rs11209948 | G | T | 0.35 | 44 (11) | 31 (11) | 71 (11) | 4 (10) | 57 (11) | 18 (11) | 64 (11) | 11 (11) | RPL31P12 |
| 1 | rs11249248 | T | C | 0.47 | 64 (16) | 43 (16) | 101 (16) | 6 (14) | 85 (17) | 22 (13) | 92 (16) | 15 (15) | LOC100288240/RHCE/SDHDP7/TMEM50A |
| 1 | rs11587012 | C | A | 0.29 | 37 (10) | 18 (6) | 53 (8) | 2 (5) | 44 (9) | 11 (7) | 50 (9) | 5 (5) | LCE1D/LCE1E/LOC100289267 |
| 1 | rs158736 | C | G | 0.34 | 35 (9) | 29 (10) | 63 (10) | 1 (2) | 52 (10) | 12 (7) | 58 (10) | 6 (6) | LOC100287974 |
| 1 | rs6693105 | T | C | 0.36 | 45 (12) | 35 (13) | 77 (12) | 3 (7) | 64 (13) | 16 (10) | 68 (12) | 12 (12) | LCE3B/LCE3C |
| 1 | rs7542235 | G | A | 0.15 | 11 (3) | 6 (2) | 16 (3) | 1 (2) | 11 (2) | 6 (4) | 16 (3) | 1 (1) | CFH/CFHR1/CFHR3/LOC100289145 |
| 2 | rs7419565 | C | T | 0.41 | 45 (12) | 39 (14) | 79 (13) | 5 (12) | 70 (14) | 14 (9) | 73 (13) | 11 (11) | TUBA3E |
| 2 | rs893403 | G | A | 0.32 | 42 (11) | 26 (9) | 66 (11) | 2 (5) | 48 (10) | 20 (12) | 55 (10) | 13 (13) | LOC100288532 |
| 5 | rs10053292 | C | T | 0.10 | 6 (2) | 3 (1) | 8 (1) | 1 (2) | 5 (1) | 4 (2) | 8 (1) | 1 (1) | HMGXB3/PDE6A/RPS20P4/SLC26A2/TIGD6 |
| 5 | rs2387715 | T | A | 0.31 | 32 (8) | 25 (9) | 55 (9) | 1 (2) | 42 (8) | 15 (9) | 46 (8) | 11 (11) | BTNL3/BTNL8/LOC100128762/LOC646227 |
| 5 | rs7703761 | C | T | 0.45 | 63 (16) | 36 (13) | 97 (16) | 2 (5) | 74 (15) | 25 (15) | 90 (16) | 9 (9) | SPINK5L2 |
| 6 | rs17654108 | T | A | 0.13 | 7 (2) | 1 (0) | 8 (1) | 0 (0) | 5 (1) | 3 (2) | 7 (1) | 1 (1) | LOC260339 |
| 7 | rs2160195 | T | A | 0.33 | 30 (8) | 30 (11) | 59 (9) | 1 (2) | 41 (8) | 19 (12) | 51 (9) | 9 (9) | TRGV1/TRGV2/TRGV3/TRGV4/TRGV5 |
| 7 | rs4621754 | G | A | 0.09 | 1 (0) | 3 (1) | 4 (1) | 0 (0) | 3 (1) | 1 (1) | 3 (1) | 1 (1) | NCAPG2 |
| 7 | rs4729606 | C | T | 0.26 | 20 (5) | 17 (6) | 35 (6) | 2 (5) | 30 (6) | 7 (4) | 32 (6) | 5 (5) | ZAN |
| 7 | rs6943474 | G | A | 0.47 | 86 (22) | 59 (21) | 134 (21) | 11 (26) | 113 (22) | 32 (20) | 125 (22) | 20 (21) | EMID2 |
| 8 | rs11985201 | A | G | 0.40 | 44 (11) | 43 (16) | 78 (13) | 9 (21) | 70 (14) | 17 (10) | 73 (13) | 14 (14) | ADAM3A/ADAM5P |

| | | | | | | | | | | | | | |
|---|---|---|---|---|---|---|---|---|---|---|---|---|---|
| 8 | rs4543566 | G | C | 0.11 | 3 (1) | 2 (1) | 4 (1) | 1 (2) | 3 (1) | 2 (1) | 4 (1) | 1 (1) | DEFA10P |
| 9 | rs1523688 | G | T | 0.24 | 24 (6) | 5 (2) | 26 (4) | 3 (7) | 22 (4) | 7 (4) | 26 (5) | 3 (3) | OR13C2/OR13C5 |
| 9 | rs2174926 | A | G | 0.44 | 58 (15) | 43 (16) | 90 (14) | 11 (26) | 71 (14) | 30 (18) | 85 (15) | 16 (16) | LOC442434 |
| 10 | rs10885336 | A | G | 0.38 | 43 (11) | 39 (14) | 78 (13) | 4 (10) | 60 (12) | 22 (13) | 68 (12) | 14 (14) | GUCY2G |
| 10 | rs2342606 | T | C | 0.44 | 51 (13) | 54 (19) | 103 (17) | 2 (5) | 82 (16) | 23 (14) | 85 (15) | 20 (20) | LOC642521/LOC642538 |
| 10 | rs3793917 | G | C | 0.25 | 14 (4) | 15 (5) | 25 (4) | 4 (10) | 21 (4) | 8 (5) | 25 (4) | 4 (4) | ARMS2 |
| 11 | rs11228868 | T | C | 0.07 | 2 (1) | 1 (0) | 3 (0) | 0 (0) | 1 (0) | 2 (1) | 3 (1) | 0 (0) | TRIM48 |
| 11 | rs1944862 | A | G | 0.30 | 23 (6) | 26 (9) | 47 (8) | 2 (5) | 37 (7) | 12 (7) | 34 (6) | 15 (15) | OR4P1P |
| 11 | rs4882017 | A | G | 0.33 | 88 (23) | 72 (26) | 151 (24) | 9 (21) | 119 (24) | 41 (25) | 139 (24) | 21 (22) | OR4A45P |
| 12 | rs1478309 | T | G | 0.18 | 14 (4) | 7 (3) | 20 (3) | 1 (2) | 16 (3) | 5 (3) | 18 (3) | 3 (3) | KLRC1/KLRC2/KLRC3 |
| 13 | rs9318648 | A | G | 0.29 | 28 (7) | 18 (6) | 43 (7) | 3 (7) | 32 (6) | 14 (9) | 42 (7) | 4 (4) | LOC374491 |
| 14 | rs11156875 | G | A | 0.19 | 12 (3) | 7 (3) | 17 (3) | 2 (5) | 14 (3) | 5 (3) | 14 (2) | 5 (5) | RPL23AP70 |
| 14 | rs8007442 | T | C | 0.42 | 59 (15) | 33 (12) | 86 (14) | 6 (14) | 68 (14) | 24 (15) | 83 (15) | 9 (9) | TRAV14DV4 |
| 14 | rs8022070 | T | C | 0.11 | 2 (1) | 3 (1) | 5 (1) | 0 (0) | 4 (1) | 1 (1) | 5 (1) | 0 (0) | LOC731308 |
| 16 | rs10521145 | A | G | 0.09 | 2 (1) | 0 (0) | 2 (0) | 0 (0) | 1 (0) | 1 (1) | 2 (0) | 0 (0) | SULT1A1 |
| 16 | rs2244613 | G | T | 0.20 | 17 (4) | 14 (5) | 29 (5) | 2 (5) | 28 (6) | 3 (2) | 27 (5) | 4 (4) | CES4 |
| 17 | rs16966699 | G | C | 0.12 | 7 (2) | 3 (1) | 10 (2) | 0 (0) | 8 (2) | 2 (1) | 10 (2) | 0 (0) | KRT33A, KRT33B |
| 17 | rs8064493 | A | G | 0.21 | 22 (6) | 9 (3) | 30 (5) | 1 (2) | 26 (5) | 5 (3) | 30 (5) | 1 (1) | KRTAP9P1 |
| 19 | rs103294 | T | C | 0.29 | 35 (9) | 19 (7) | 50 (8) | 4 (10) | 30 (8) | 16 (10) | 47 (8) | 7 (7) | LILRA3 |
| 19 | rs324121 | A | G | 0.13 | 8 (2) | 7 (3) | 14 (2) | 1 (2) | 8 (2) | 7 (4) | 12 (2) | 3 (3) | LOC400713 |
| 19 | rs3810336 | A | G | 0.28 | 31 (8) | 19 (7) | 50 (8) | 0 (0) | 38 (8) | 12 (7) | 46 (8) | 4 (4) | GALP |
| 19 | rs4806152 | C | A | 0.23 | 22 (6) | 13 (5) | 34 (5) | 1 (2) | 29 (6) | 6 (4) | 31 (5) | 4 (4) | FFAR3/GPR42P |

A1, minor allele; A2, major allele; CHR, chromosome; MAF, minor allele frequency

[a]Mismatch refers to cases in which a recipient who is homozygous for a deletion-tagging allele received a transplant from a donor with a nonhomozygous or homozygous reference allele genotype.

[b]Steers NJ, Li Y, Drace Z, et al. Genomic Mismatch at LIMS1 Locus and Kidney Allograft Rejection. *N Engl J Med*. 2019;380(20):1918-1928. doi:10.1056/nejmoa180373

**Supplementary Table S2. Summary statistics of adjusted Cox proportional hazards models analyzing the effect of mismatches in 40 deletion-tagging variants and time to acute rejection**

| Deletion-tagging variant | Adjusted Cox proportional hazards models[a] | | FDR |
|---|---|---|---|
| | HR (95% CI) | P-value | |
| rs10927864 | 0.829(0.489–1.406) | 0.486 | 0.881 |
| rs11209948 | 0.891(0.609–1.304) | 0.552 | 0.881 |
| rs11249248 | 0.907(0.653–1.261) | 0.561 | 0.881 |
| rs11587012 | 0.744(0.459–1.208) | 0.232 | 0.619 |
| rs158736 | 1.380(0.929–2.049) | 0.111 | 0.504 |
| rs6693105 | 1.085(0.759–1.552) | 0.654 | 0.881 |
| rs7542235 | 0.598(0.264–1.352) | 0.217 | 0.619 |
| rs7419565 | 1.195(0.843–1.693) | 0.317 | 0.669 |
| rs893403 | 0.896(0.591–1.356) | 0.602 | 0.881 |
| rs10053292 | 0.916(0.289–2.906) | 0.882 | 0.954 |
| rs2387715 | 1.240(0.816–1.885) | 0.313 | 0.669 |
| rs7703761 | 0.754(0.525–1.082) | 0.126 | 0.504 |
| rs17654108 | 0.177(0.025–1.271) | 0.085 | 0.504 |
| rs2160195 | 1.214(0.829–1.778) | 0.318 | 0.669 |
| rs4621754 | 2.533(0.798–8.041) | 0.115 | 0.504 |
| rs4729606 | 1.139(0.684–1.897) | 0.618 | 0.881 |
| rs11985201 | 1.483(1.066–2.062) | 0.019 | 0.448 |
| rs4543566 | 0.878(0.215–3.580) | 0.856 | 0.951 |
| rs1523688 | 0.419(0.172–1.021) | 0.056 | 0.448 |
| rs2174926 | 0.989(0.709–1.380) | 0.948 | 0.972 |
| rs10885336 | 1.093(0.776–1.540) | 0.610 | 0.881 |
| rs2342606 | 1.373(1.011–1.865) | 0.042 | 0.448 |
| rs3793917 | 1.328(0.783–2.254) | 0.292 | 0.669 |
| rs11228868 | 1.216(0.169–8.755) | 0.846 | 0.951 |
| rs1944862 | 1.355(0.897–2.048) | 0.149 | 0.542 |
| rs1478309 | 0.611(0.287–1.302) | 0.201 | 0.618 |
| rs9318648 | 0.927(0.572–1.504) | 0.760 | 0.950 |
| rs11156875 | 1.175(0.519–2.661) | 0.699 | 0.902 |
| rs8007442 | 0.779(0.535–1.135) | 0.193 | 0.618 |
| rs8022070 | 2.505(0.787–7.975) | 0.120 | 0.504 |
| rs10521145 | 0(0–Inf) | 0.991 | 0.991 |
| rs2244613 | 1.063(0.619–1.828) | 0.824 | 0.951 |
| rs16966699 | 0.674(0.214–2.120) | 0.499 | 0.881 |
| rs8064493 | 0.513(0.260–1.012) | 0.054 | 0.448 |
| rs103294 | 0.900(0.564–1.438) | 0.661 | 0.881 |
| rs324121 | 1.105(0.516–2.367) | 0.797 | 0.951 |
| rs3810336 | 0.982(0.613–1.571) | 0.938 | 0.972 |
| rs4806152 | 0.780(0.442–1.378) | 0.392 | 0.784 |
| rs6943474 | 0.914(0.681–1.227) | 0.550 | 0.881 |
| rs4882017 | 1.313(1.000–1.722) | 0.050 | 0.448 |

CI, confidence interval; FDR, false detection rate; HR, hazard ratio.

[a] Models were adjusted with recipient and donor age, recipient and donor sex, cold ischemia time, HLA I eplet mismatch, HLA II eplet mismatch, year of transplantation, autoimmune status and calcineurin inhibitor type initial status.

**Supplementary Table S3. Summary statistics of adjusted Cox proportional hazards models analyzing the effect of mismatches in 40 deletion-tagging variants and time to late rejection**

| Deletion-tagging variant | Adjusted Cox proportional hazards models[a] | | FDR |
|---|---|---|---|
| | HR (95% CI) | P-value | |
| rs10927864 | 1.004(0.434–2.326) | 0.992 | 0.995 |
| rs11209948 | 0.967(0.513–1.823) | 0.916 | 0.995 |
| rs11249248 | 0.903(0.516–1.582) | 0.722 | 0.995 |
| rs11587012 | 0.625(0.252–1.552) | 0.311 | 0.982 |
| rs158736 | 0.673(0.291–1.556) | 0.355 | 0.995 |
| rs6693105 | 1.003(0.544–1.847) | 0.993 | 0.995 |
| rs7542235 | 0.322(0.045–2.328) | 0.262 | 0.953 |
| rs7419565 | 0.869(0.463–1.633) | 0.663 | 0.995 |
| rs893403 | 1.504(0.831–2.721) | 0.178 | 0.953 |
| rs10053292 | 1.025(0.140–7.524) | 0.980 | 0.995 |
| rs2387715 | 1.517(0.803–2.864) | 0.199 | 0.953 |
| rs7703761 | 0.608(0.304–1.214) | 0.158 | 0.953 |
| rs17654108 | 0.807(0.110–5.925) | 0.833 | 0.995 |
| rs2160195 | 1.039(0.522–2.069) | 0.913 | 0.995 |
| rs4621754 | 1.946(0.264–14.358) | 0.514 | 0.995 |
| rs4729606 | 1.025(0.413–2.545) | 0.958 | 0.995 |
| rs11985201 | 1.196(0.676–2.114) | 0.538 | 0.995 |
| rs4543566 | 1.439(0.196–10.554) | 0.720 | 0.995 |
| rs1523688 | 0.864(0.270–2.769) | 0.806 | 0.995 |
| rs2174926 | 1.173(0.681–2.020) | 0.566 | 0.995 |
| rs10885336 | 1.205(0.681–2.130) | 0.522 | 0.995 |
| rs2342606 | 1.530(0.926–2.528) | 0.097 | 0.953 |
| rs3793917 | 0.867(0.316–2.378) | 0.782 | 0.995 |
| rs11228868 | 0(0–Inf) | 0.994 | 0.995 |
| rs1944862 | 2.341(1.326–4.130) | 0.003 | 0.120 |
| rs1478309 | 0.930(0.291–2.964) | 0.902 | 0.995 |
| rs9318648 | 0.553(0.201–1.521) | 0.251 | 0.953 |
| rs11156875 | 2.558(0.925–7.074) | 0.070 | 0.933 |
| rs8007442 | 0.654(0.327–1.308) | 0.230 | 0.953 |
| rs8022070 | 0(0–Inf) | 0.994 | 0.995 |
| rs10521145 | 0(0–Inf) | 0.995 | 0.995 |
| rs2244613 | 0.820(0.299–2.248) | 0.700 | 0.995 |
| rs16966699 | 0(0–Inf) | 0.995 | 0.995 |
| rs8064493 | 0.145(0.020–1.057) | 0.057 | 0.933 |
| rs103294 | 0.954(0.441–2.065) | 0.905 | 0.995 |
| rs324121 | 1.179(0.365–3.810) | 0.784 | 0.995 |
| rs3810336 | 0.533(0.195–1.457) | 0.220 | 0.953 |
| rs4806152 | 0.595(0.215–1.652) | 0.319 | 0.982 |
| rs6943474 | 0.891(0.538–1.476) | 0.654 | 0.995 |
| rs4882017 | 0.946(0.581–1.540) | 0.823 | 0.995 |

CI, confidence interval; FDR, false detection rate; HR, hazard ratio.

[a] Models were adjusted with recipient and donor age, recipient and donor sex, cold ischemia time, HLA I eplet mismatch, HLA II eplet mismatch, year of transplantation, autoimmune status and calcineurin inhibitor type initial status.

**Supplementary Table S4. Summary statistics of adjusted Cox proportional hazards models analyzing the effect of mismatches in 40 deletion-tagging variants and time to graft loss**

| Deletion-tagging variant | Adjusted Cox proportional hazards models[a] | | FDR |
|---|---|---|---|
| | HR (95% CI) | P-value | |
| rs10927864 | 0.873(0.203–3.750) | 0.855 | 0.997 |
| rs11209948 | 0.839(0.294–2.390) | 0.742 | 0.997 |
| rs11249248 | 0.771(0.320–1.855) | 0.561 | 0.997 |
| rs11587012 | 0.484(0.114–2.051) | 0.325 | 0.997 |
| rs158736 | 0.313(0.042–2.316) | 0.255 | 0.997 |
| rs6693105 | 0.598(0.183–1.960) | 0.396 | 0.997 |
| rs7542235 | 0.919(0.122–6.912) | 0.935 | 0.997 |
| rs7419565 | 1.030(0.399–2.658) | 0.951 | 0.997 |
| rs893403 | 0.492(0.117–2.061) | 0.332 | 0.997 |
| rs10053292 | 3.760(0.478–29.554) | 0.208 | 0.997 |
| rs2387715 | 0.639(0.151–2.698) | 0.543 | 0.997 |
| rs7703761 | 0.298(0.071–1.249) | 0.098 | 0.980 |
| rs17654108 | 0(0–Inf) | 0.996 | 0.997 |
| rs2160195 | 0.265(0.036–1.940) | 0.191 | 0.997 |
| rs4621754 | 0(0–Inf) | 0.997 | 0.997 |
| rs4729606 | 1.110(0.262–4.705) | 0.887 | 0.997 |
| rs11985201 | 1.621(0.739–3.558) | 0.228 | 0.997 |
| rs4543566 | 3.970(0.510–30.902) | 0.188 | 0.997 |
| rs1523688 | 2.601(0.760–8.901) | 0.128 | 0.997 |
| rs2174926 | 2.332(1.145–4.752) | 0.020 | 0.800 |
| rs10885336 | 0.811(0.286–2.297) | 0.693 | 0.997 |
| rs2342606 | 0.290(0.069–1.212) | 0.090 | 0.98 |
| rs3793917 | 2.538(0.874–7.372) | 0.087 | 0.980 |
| rs11228868 | 0(0–Inf) | 0.996 | 0.997 |
| rs1944862 | 0.686(0.160–2.946) | 0.613 | 0.997 |
| rs1478309 | 0.839(0.114–6.182) | 0.863 | 0.997 |
| rs9318648 | 1.070(0.322–3.555) | 0.912 | 0.997 |
| rs11156875 | 1.299(0.174–9.703) | 0.799 | 0.997 |
| rs8007442 | 1.087(0.444–2.661) | 0.854 | 0.997 |
| rs8022070 | 0(0–Inf) | 0.996 | 0.997 |
| rs10521145 | 0(0–Inf) | 0.997 | 0.997 |
| rs2244613 | 0.985(0.235–4.130) | 0.983 | 0.997 |
| rs16966699 | 0(0–Inf) | 0.996 | 0.997 |
| rs8064493 | 0.535(0.071–4.021) | 0.544 | 0.997 |
| rs103294 | 1.359(0.478–3.865) | 0.565 | 0.997 |
| rs324121 | 0.704(0.094–5.301) | 0.734 | 0.997 |
| rs3810336 | 0(0–Inf) | 0.995 | 0.997 |
| rs4806152 | 0.469(0.063–3.491) | 0.460 | 0.997 |
| rs6943474 | 0.968(0.457–2.050) | 0.932 | 0.997 |
| rs4882017 | 1.061(0.496–2.267) | 0.879 | 0.997 |

CI, confidence interval; FDR, false detection rate; HR, hazard ratio.

[a] Models were adjusted with recipient and donor age, recipient and donor sex, cold ischemia time, HLA I eplet mismatch, HLA II eplet mismatch, year of transplantation, autoimmune status and calcineurin inhibitor type initial status.

**Supplementary Table S5. Summary statistics of adjusted Cox proportional hazards models analyzing the effect of mismatches in 40 deletion-tagging variants and time to death.**

| Deletion-tagging variant | Adjusted Cox proportional hazards models[a] | | FDR |
|---|---|---|---|
| | HR (95% CI) | P-value | |
| rs10927864 | 0.899(0.469–1.722) | 0.748 | 0.938 |
| rs11209948 | 0.806(0.478–1.361) | 0.420 | 0.832 |
| rs11249248 | 0.764(0.475–1.230) | 0.268 | 0.814 |
| rs11587012 | 0.792(0.425–1.477) | 0.464 | 0.844 |
| rs158736 | 0.703(0.367–1.344) | 0.286 | 0.814 |
| rs6693105 | 0.759(0.444–1.299) | 0.315 | 0.814 |
| rs7542235 | 1.518(0.660–3.490) | 0.326 | 0.814 |
| rs7419565 | 0.725(0.416–1.266) | 0.258 | 0.814 |
| rs893403 | 1.232(0.756–2.007) | 0.402 | 0.832 |
| rs10053292 | 1.748(0.546–5.593) | 0.346 | 0.814 |
| rs2387715 | 1.238(0.723–2.120) | 0.437 | 0.832 |
| rs7703761 | 1.151(0.743–1.785) | 0.529 | 0.920 |
| rs17654108 | 4.271(1.303–13.999) | 0.017 | 0.340 |
| rs2160195 | 1.487(0.917–2.411) | 0.108 | 0.814 |
| rs4621754 | 1.162(0.160–8.468) | 0.882 | 0.980 |
| rs4729606 | 0.858(0.399–1.844) | 0.695 | 0.938 |
| rs11985201 | 0.741(0.440–1.247) | 0.259 | 0.814 |
| rs4543566 | 1.272(0.175–9.248) | 0.812 | 0.955 |
| rs1523688 | 1.135(0.524–2.457) | 0.748 | 0.938 |
| rs2174926 | 1.329(0.887–1.990) | 0.168 | 0.814 |
| rs10885336 | 1.294(0.820–2.042) | 0.268 | 0.814 |
| rs2342606 | 0.724(0.448–1.170) | 0.187 | 0.814 |
| rs3793917 | 1.133(0.552–2.327) | 0.734 | 0.938 |
| rs11228868 | 7.554(1.767–32.295) | 0.006 | 0.240 |
| rs1944862 | 1.010(0.542–1.883) | 0.975 | 0.999 |
| rs1478309 | 1.027(0.420–2.514) | 0.953 | 0.999 |
| rs9318648 | 1.030(0.577–1.840) | 0.921 | 0.996 |
| rs11156875 | 1.273(0.517–3.136) | 0.600 | 0.935 |
| rs8007442 | 0.966(0.613–1.523) | 0.882 | 0.980 |
| rs8022070 | 1.001(0.137–7.321) | 0.999 | 0.999 |
| rs10521145 | 5.276(0.714–38.984) | 0.103 | 0.814 |
| rs2244613 | 0.406(0.129–1.274) | 0.122 | 0.814 |
| rs16966699 | 1.413(0.345–5.798) | 0.631 | 0.935 |
| rs8064493 | 0.624(0.252–1.543) | 0.307 | 0.814 |
| rs103294 | 1.252(0.733–2.138) | 0.411 | 0.832 |
| rs324121 | 1.791(0.825–3.888) | 0.141 | 0.814 |
| rs3810336 | 1.091(0.602–1.978) | 0.774 | 0.938 |
| rs4806152 | 0.799(0.348–1.833) | 0.597 | 0.935 |
| rs6943474 | 0.940(0.632–1.398) | 0.760 | 0.938 |
| rs4882017 | 1.095(0.759–1.579) | 0.629 | 0.935 |

CI, confidence interval; FDR, false detection rate; HR, hazard ratio.

[a] Models were adjusted with recipient and donor age, recipient and donor sex, cold ischemia time, HLA I eplet mismatch, HLA II eplet mismatch, year of transplantation, autoimmune status and calcineurin inhibitor type initial status.

**Supplementary Table S6. Adjusted Cox proportional hazards model for rs11985201 mismatch and time to acute rejection.**

| Covariate | HR (95% CI) | P-value |
|---|---|---|
| rs11985201 mismatch | 1.483(1.066–2.062) | 0.019 |
| Recipient age | 0.982(0.973–0.992) | 0.001 |
| Donor age | 0.997(0.988–1.006) | 0.495 |
| Recipient sex | 1.140(0.887–1.467) | 0.307 |
| Donor sex | 1.172(0.912–1.506) | 0.214 |
| Cold ischemia time minutes | 0.999(0.998–1.000) | 0.127 |
| Eplets total HLAI | 1.019(1.002–1.036) | 0.027 |
| Eplets total HLAII | 1.018(0.995–1.042) | 0.123 |
| Year of transplantation | 0.966(0.943–0.990) | 0.007 |
| Autoimmune liver disease | 1.260(0.978–1.623) | 0.074 |
| Initial calcineurin inhibitor type: cyclosporine A | 1.585(1.124–2.235) | 0.009 |

Autoimmune liver disease was categorized as primary or secondary liver transplantation indication of primary sclerosing cholangitis, primary biliary cholangitis, or autoimmune hepatitis. CI; confidence interval, HR; hazard ration, HLA; human leukocyte antigen.

**Supplementary Table S7. Adjusted Cox proportional hazards model for rs2342606 mismatch and time to acute rejection.**

| Covariate | HR (95% CI) | P-value |
|---|---|---|
| rs2342606 mismatch | 1.373(1.011–1.865) | 0.042 |
| Recipient age | 0.982(0.972–0.992) | <0.001 |
| Donor age | 0.997(0.988–1.006) | 0.497 |
| Recipient sex | 1.131(1.879–1.455) | 0.340 |
| Donor sex | 1.159(1.901–1.489) | 0.250 |
| Cold ischemia time minutes | 0.999(0.998–1.000) | 0.164 |
| Eplets total HLAI | 0.016(0.000–1.033) | 0.057 |
| Eplets total HLAII | 0.021(0.997–1.045) | 0.081 |
| Year of transplantation | 0.965(0.942–0.989) | 0.005 |
| Autoimmune liver disease | 1.210(1.939–1.561) | 0.141 |
| Initial calcineurin inhibitor type: cyclosporine A | 1.635(1.158–2.308) | 0.005 |

Autoimmune liver disease was categorized as primary or secondary liver transplantation indication of primary sclerosing cholangitis, primary biliary cholangitis, or autoimmune hepatitis.CI; confidence interval, HR; hazard ration, HLA; human leukocyte antigen.

**Supplementary Table S8. Adjusted Cox proportional hazards model for rs1944862 mismatch and time to late rejection.**

| Covariate | HR (95% CI) | P-value |
|---|---|---|
| rs1944862 mismatch | 2.341(1.326–4.130) | 0.003 |
| Recipient age | 0.990(0.973–1.006) | 0.225 |
| Donor age | 0.993(0.979–1.008) | 0.371 |
| Recipient sex | 1.318(0.862–2.014) | 0.202 |
| Donor sex | 1.035(0.674–1.590) | 0.875 |
| Cold ischemia time minutes | 1.000(0.998–1.002) | 0.767 |
| Eplets total HLAI | 1.010(0.981–1.039) | 0.512 |
| Eplets total HLAII | 1.032(0.992–1.074) | 0.121 |
| Year of transplantation | 0.983(0.942–1.026) | 0.443 |
| Autoimmune liver disease | 1.087(0.705–1.675) | 0.707 |
| Initial calcineurin inhibitor type: cyclosporine A | 2.151(1.128–4.100) | 0.020 |

Autoimmune liver disease was categorized as primary or secondary liver transplantation indication of primary sclerosing cholangitis, primary biliary cholangitis, or autoimmune hepatitis. CI; confidence interval, HR; hazard ration, HLA; human leukocyte antigen.

**Supplementary Table S9. Adjusted Cox proportional hazards model for rs2174926 mismatch and time to graft loss.**

| Covariate | HR (95% CI) | P-value |
|---|---|---|
| rs2174926 mismatch | 2.332(1.145–4.752) | 0.020 |
| Recipient age | 0.972(0.947–0.998) | 0.034 |
| Donor age | 1.030(1.002–1.059) | 0.037 |
| Recipient sex | 1.157(0.591–2.263) | 0.671 |
| Donor sex | 1.123(0.569–2.218) | 0.738 |
| Cold ischemia time minutes | 1.000(0.997–1.003) | 0.971 |
| Eplets total HLAI | 0.972(0.928–1.017) | 0.215 |
| Eplets total HLAII | 1.037(0.978–1.101) | 0.227 |
| Year of transplantation | 0.959(0.890–1.033) | 0.266 |
| Autoimmune liver disease | 1.941(1.019–3.697) | 0.044 |
| Initial calcineurin inhibitor type: cyclosporine A | 5.629(1.331–23.809) | 0.019 |

Autoimmune liver disease was categorized as primary or secondary liver transplantation indication of primary sclerosing cholangitis, primary biliary cholangitis, or autoimmune hepatitis. CI; confidence interval, HR; hazard ration, HLA; human leukocyte antigen

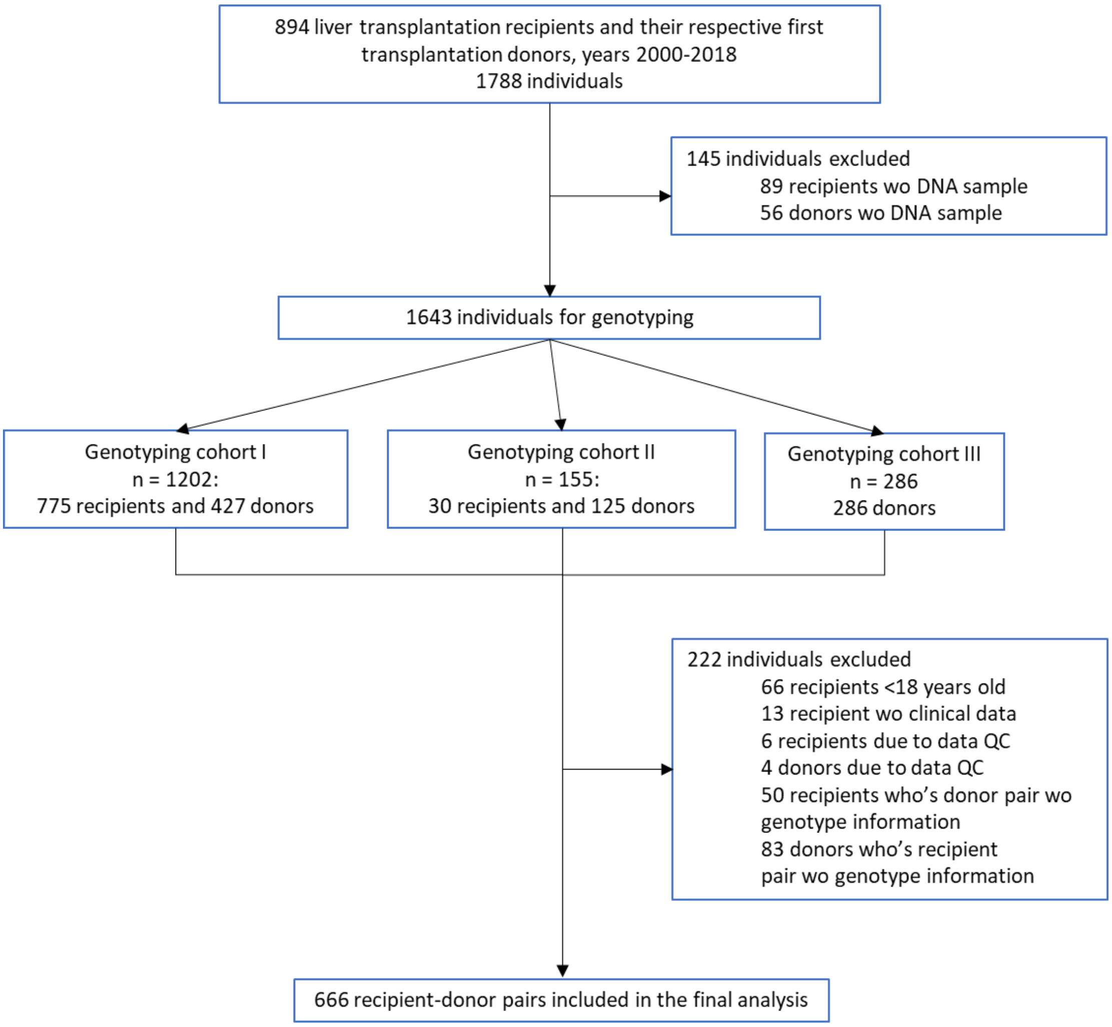

**Supplementary Figure S1. Flow of the study population. Enrolling and exclusion of the liver transplantation recipients and donors.** DNA, deoxyribonucleic acid; QC, quality control.

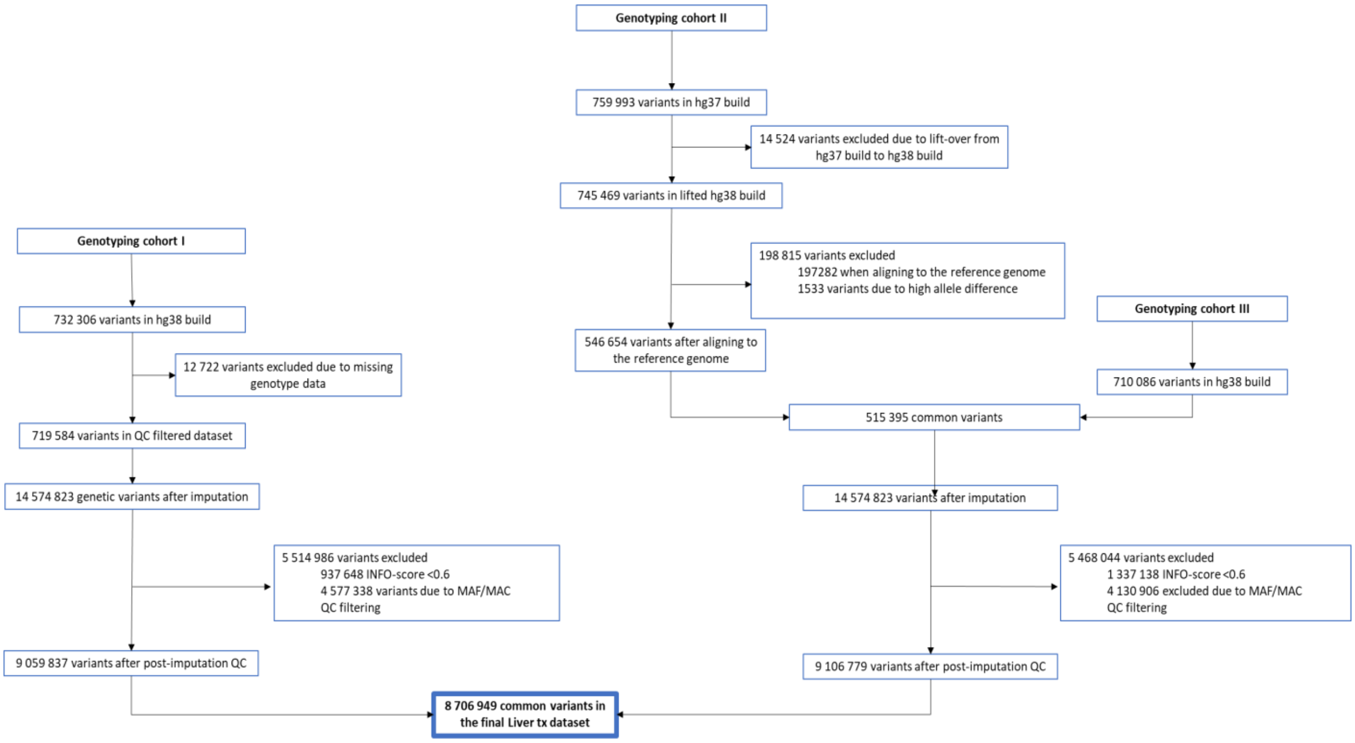

**Supplementary Figure S2. Flow of genetic variants in in pre- and post-imputation processes.** MAC, minor allele content; MAF, minor allele frequency; QC, quality control.

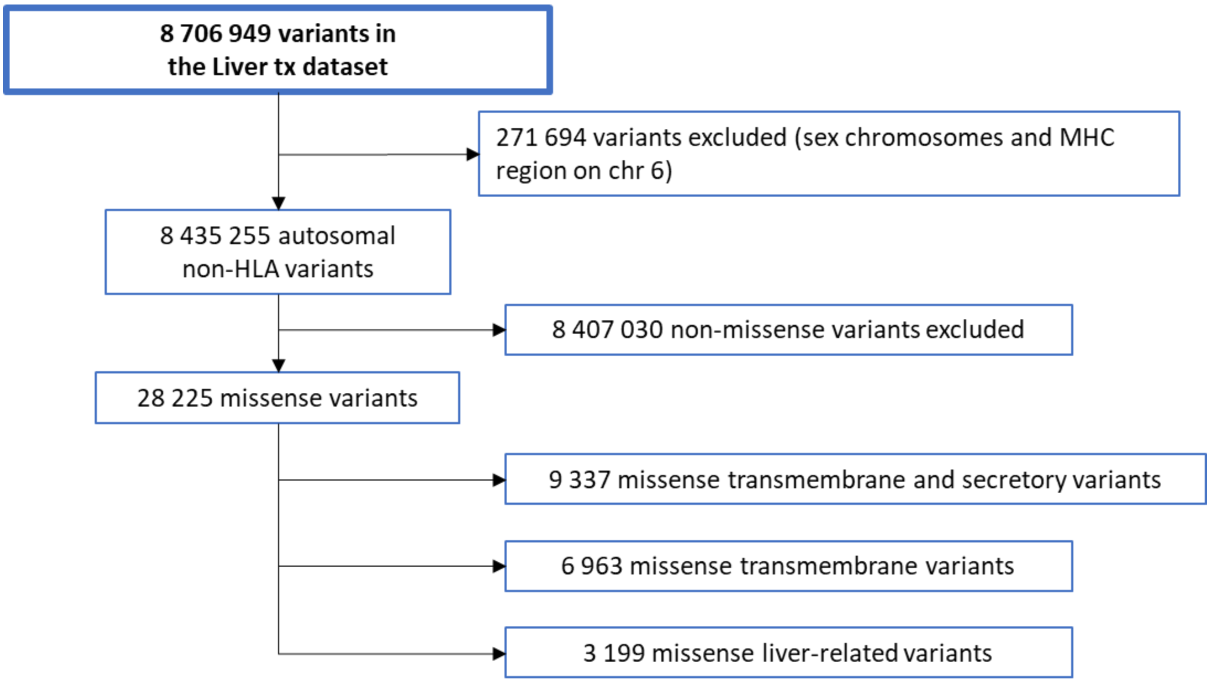

**Supplementary Figure S3. Flow of genetic variants.** HLA, human leukocyte antigen; MHC, major histocompatibility complex.

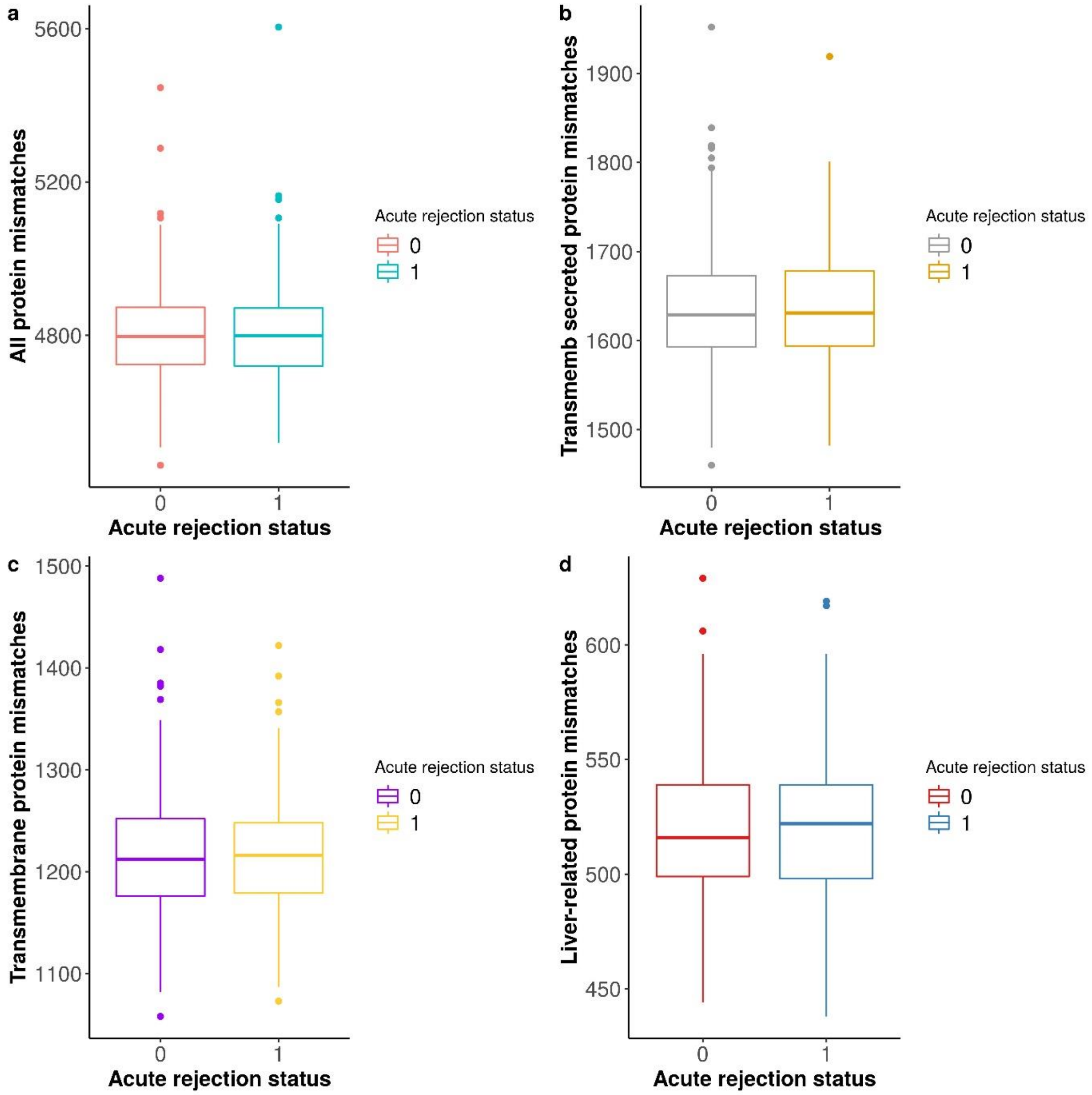

**Supplementary Figure S4. Box plots of missense variants in all four protein groups based on acute rejection status.** Distribution of missense variants coding for **(a)** all proteins, **(b)** transmembrane and secreted proteins, **(c)** transmembrane proteins, and **(d)** liver-related proteins. 0 = “no rejection” group, 1 = “rejection” group. Box plots depicts median and inter quartile range (IQR) with whiskers ±1.5 * IQR of the hinge.

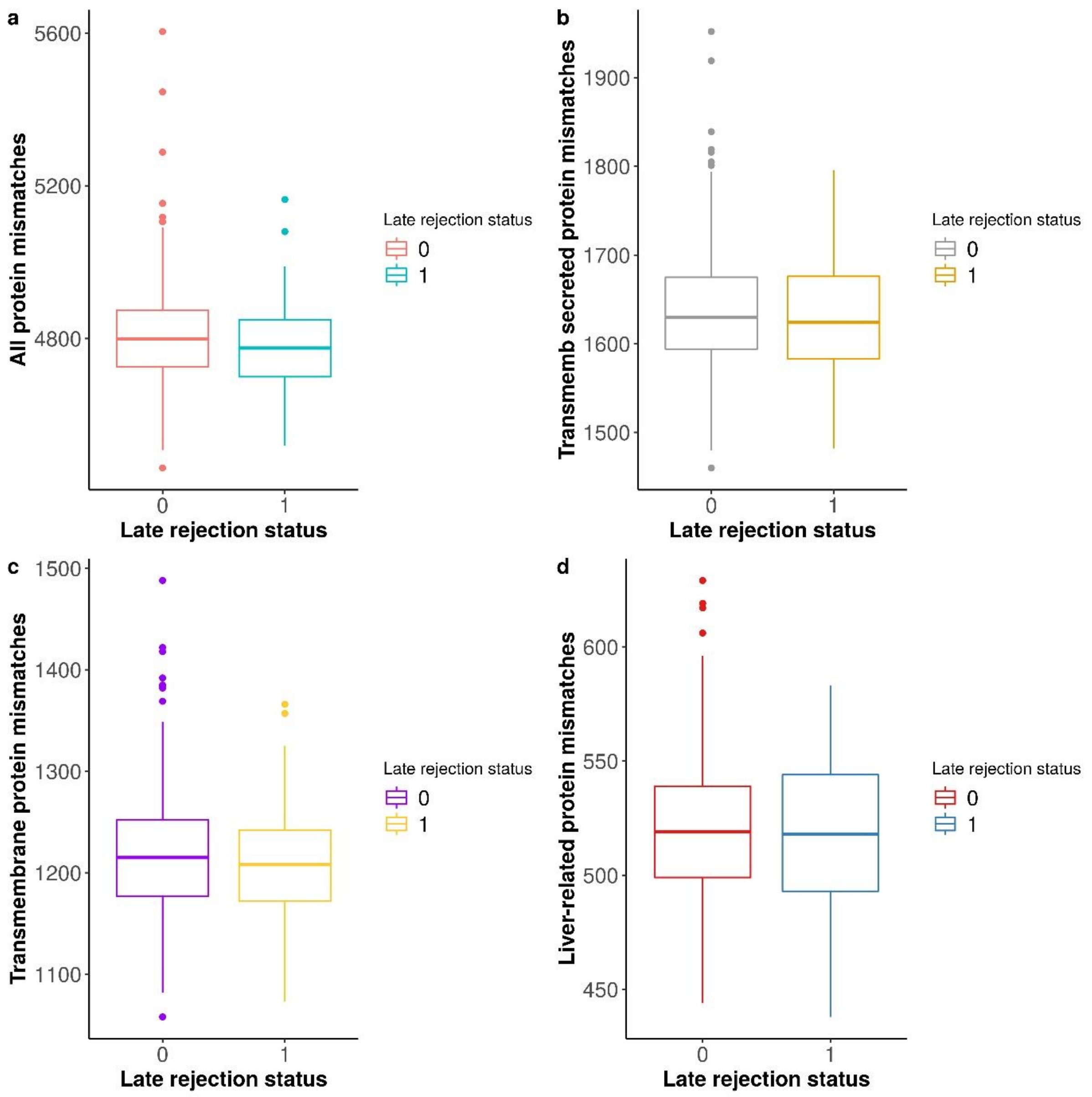

**Supplementary Figure S5. Box plots of missense variants in all four protein groups based on late rejection status.** Distribution of missense variants coding for **(a)** all proteins, **(b)** transmembrane and secreted proteins, **(c)** transmembrane proteins, and **(d)** liver-related proteins. 0 = "no late rejection" group, 1 = " late rejection" group. Box plots depicts median and inter quartile range (IQR) with whiskers ±1.5 * IQR of the hinge.

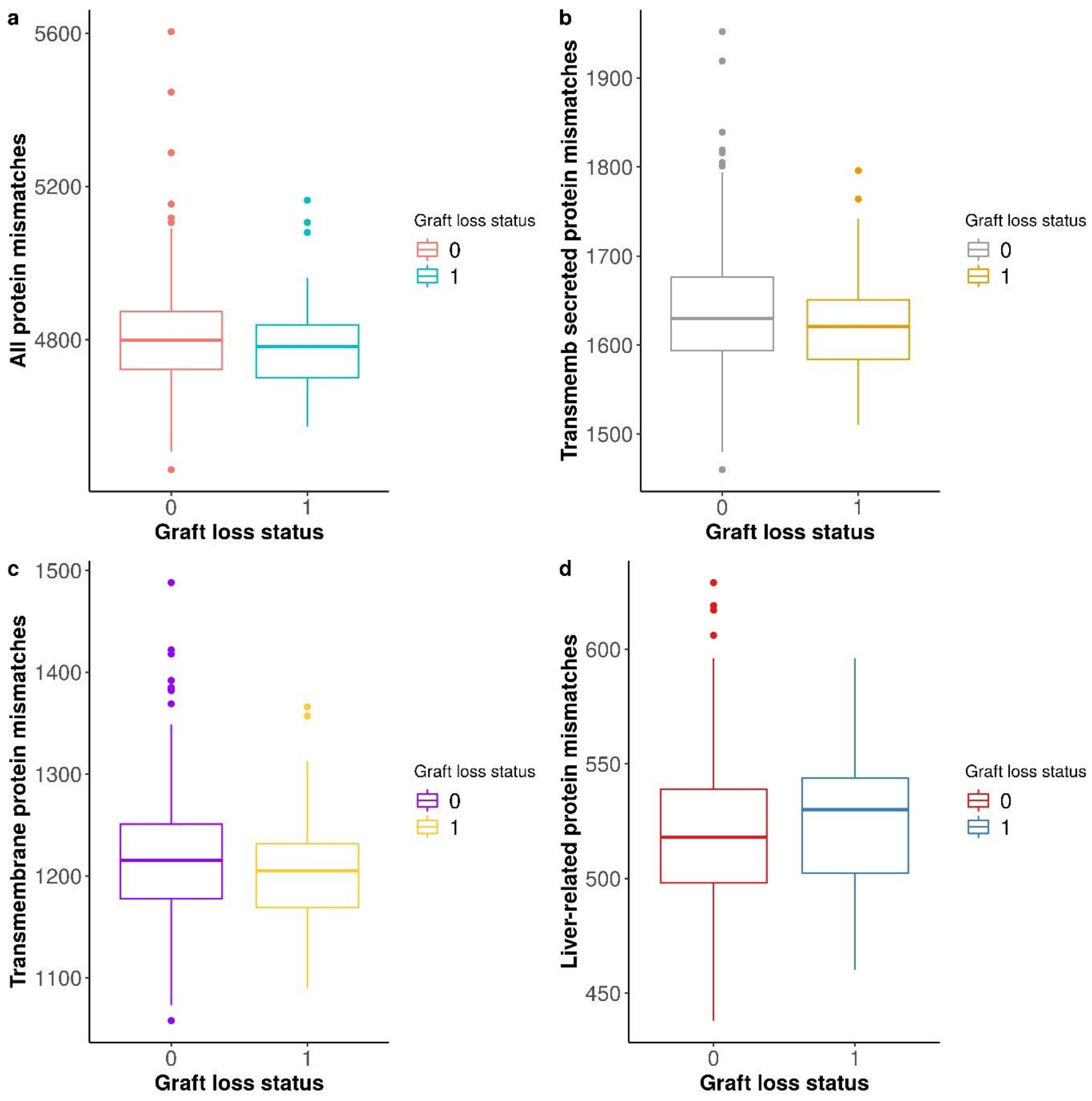

**Supplementary Figure S6. Box plots of missense variants in all four protein groups based on graft loss status.** Distribution of missense variants coding for **(a)** all proteins, **(b)** transmembrane and secreted proteins, **(c)** transmembrane proteins, and **(d)** liver-related proteins. 0 = "no graft loss" group, 1 = "graft loss" group. Box plots depicts median and inter quartile range (IQR) with whiskers ±1.5 * IQR of the hinge.

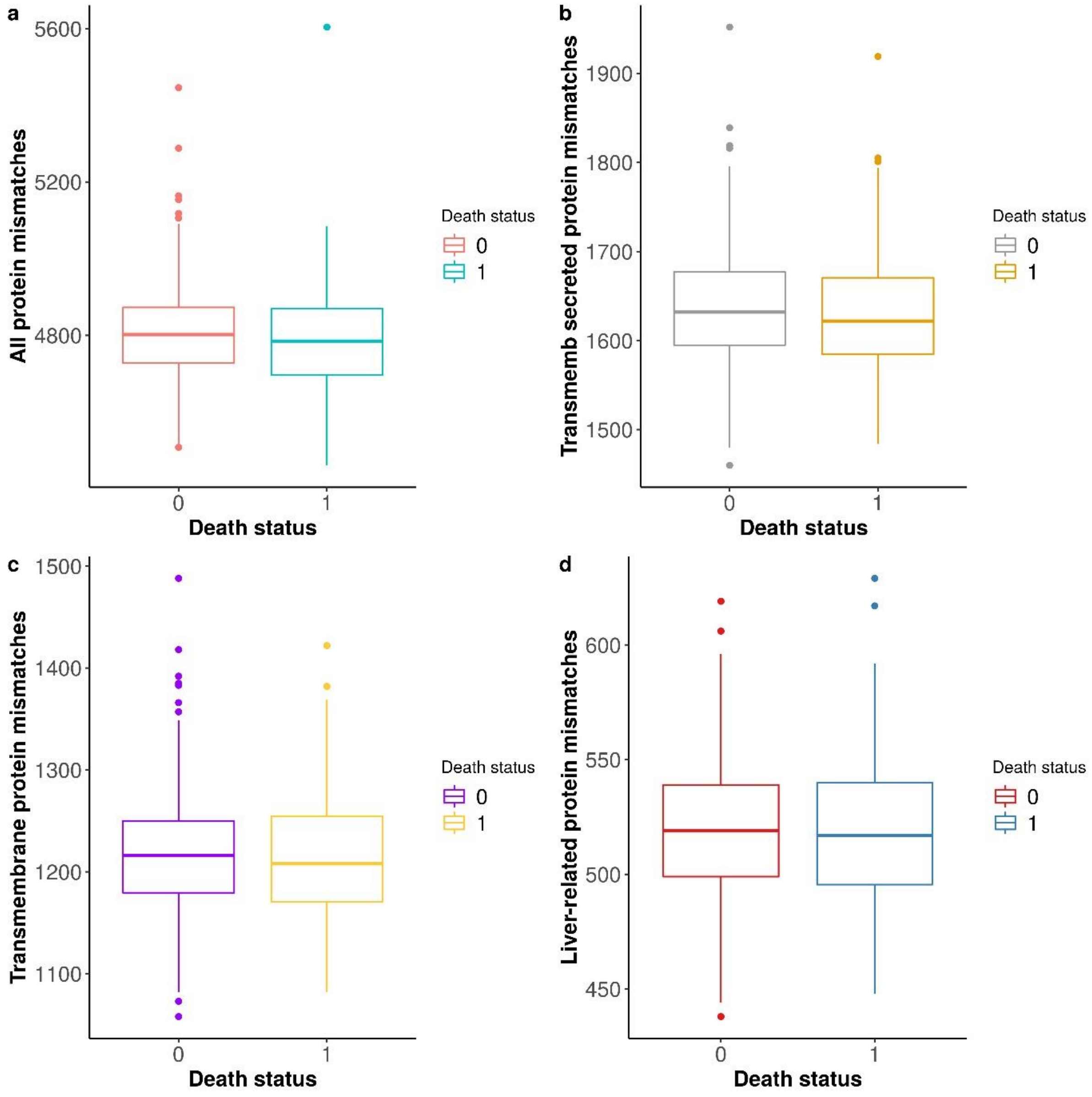

**Supplementary Figure S7. Box plots presenting the distribution of missense variants in all four protein groups based on death status.** Distribution of missense variants coding for **(a)** all proteins, **(b)** transmembrane and secreted proteins, **(c)** transmembrane proteins, and **(d)** liver-related proteins. 0 = “alive” group, 1 = “deceased” group. Box plots depicts median and inter quartile range (IQR) with whiskers ±1.5 * IQR of the hinge.

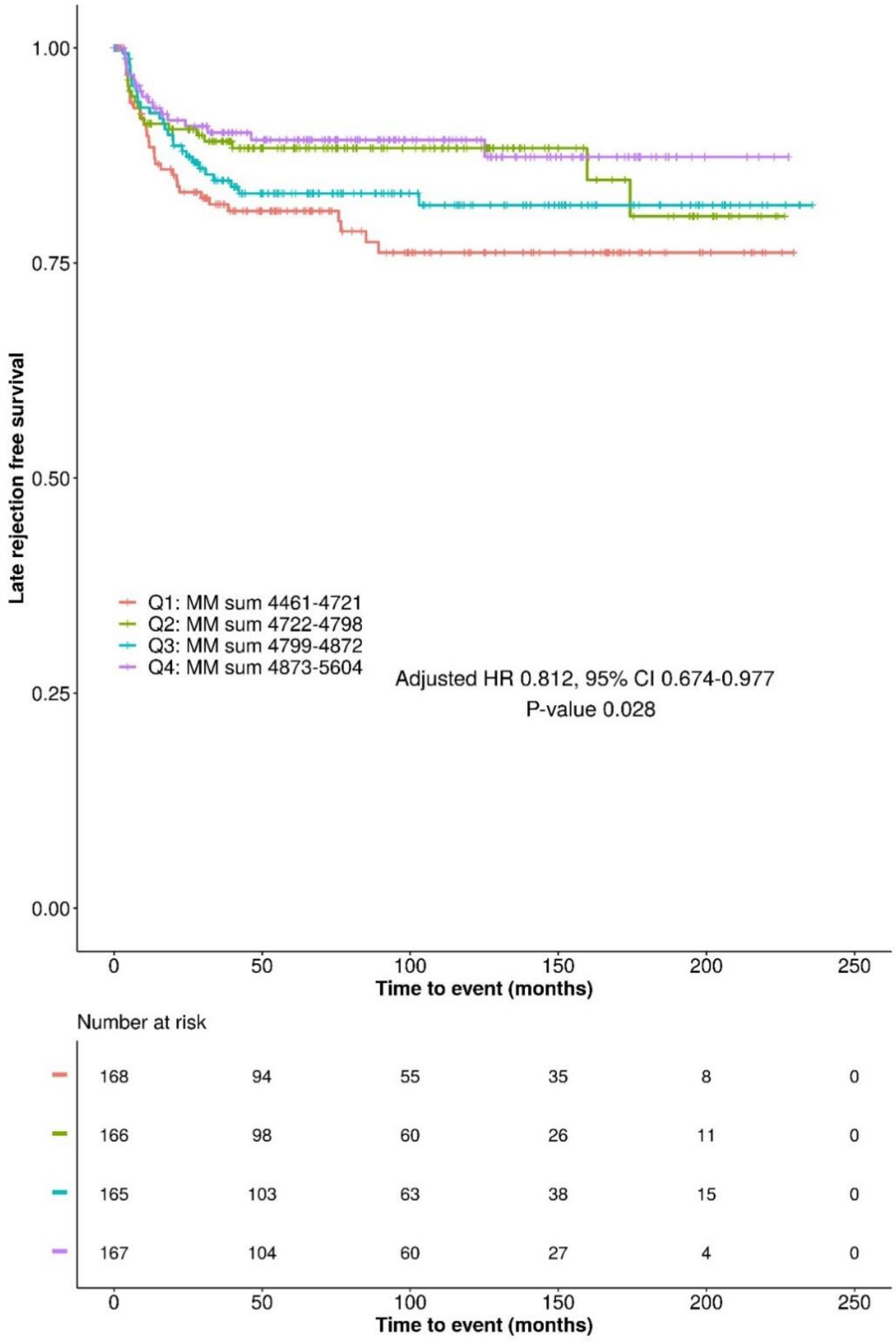

**Supplementary Figure S8. Kaplan-Meier plot displaying the effect of quartiles of missense variant mismatches of all proteins on late rejection -free survival.** The risk table shows the number of recipients in each quartile group that are at risk of late rejection at given time points. The x-axis shows time to event (late rejection or censoring) and the y-axis late rejection-free survival. The presented hazard ratio, confidence intervals and P-value were calculate using Cox proportional hazards model with adjusted covariates: recipient and donor age, recipient and donor sex, cold ischemia time, HLA I eplet mismatch, HLA II eplet mismatch, year of transplantation, autoimmune liver disease and initial calcineurin inhibitor type. CI, confidence interval; HR, hazard ratio; MM, missense variant mismatch; Q1, quartile 1; Q2, quartile 2; Q3, quartile 3; Q4, quartile 4.

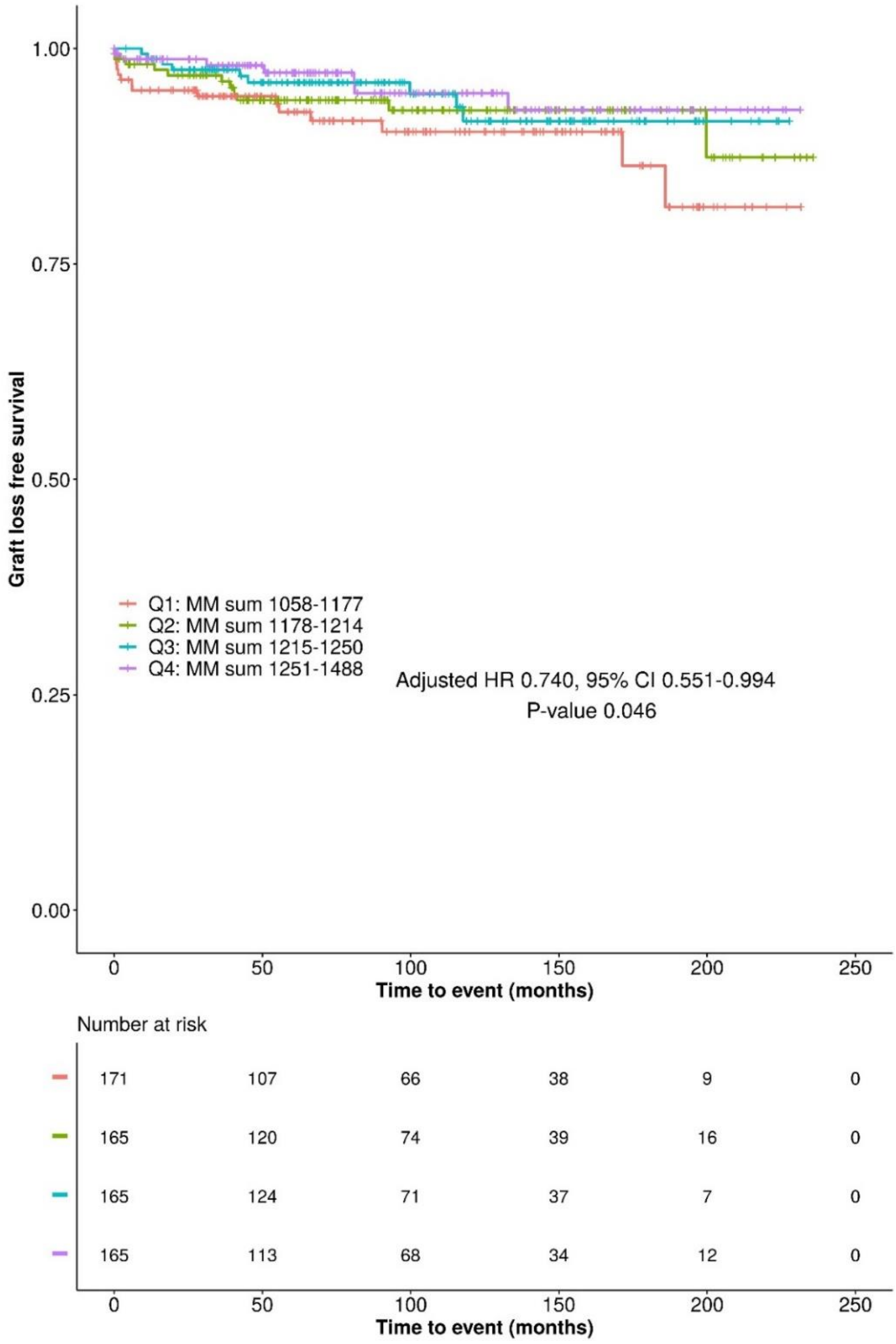

**Supplementary Figure S9. Kaplan-Meier plot displaying the effect of quartiles of missense variant mismatches of transmembrane proteins only on graft loss -free survival.** The risk table shows the number of recipients in each quartile group that are at risk of graft loss at given time points. The x-axis shows time to event (graft loss or censoring) and the y-axis graft loss-free survival. The presented hazard ratio, confidence intervals and P-value were calculate using Cox proportional hazards model with adjusted covariates: recipient and donor age, recipient and donor sex, cold ischemia time, HLA I eplet mismatch, HLA II eplet mismatch, year of transplantation, autoimmune liver disease and initial calcineurin inhibitor type. CI, confidence interval; HR, hazard ratio; MM, missense variant mismatch; Q1, quartile 1; Q2, quartile 2; Q3, quartile 3; Q4, quartile 4.

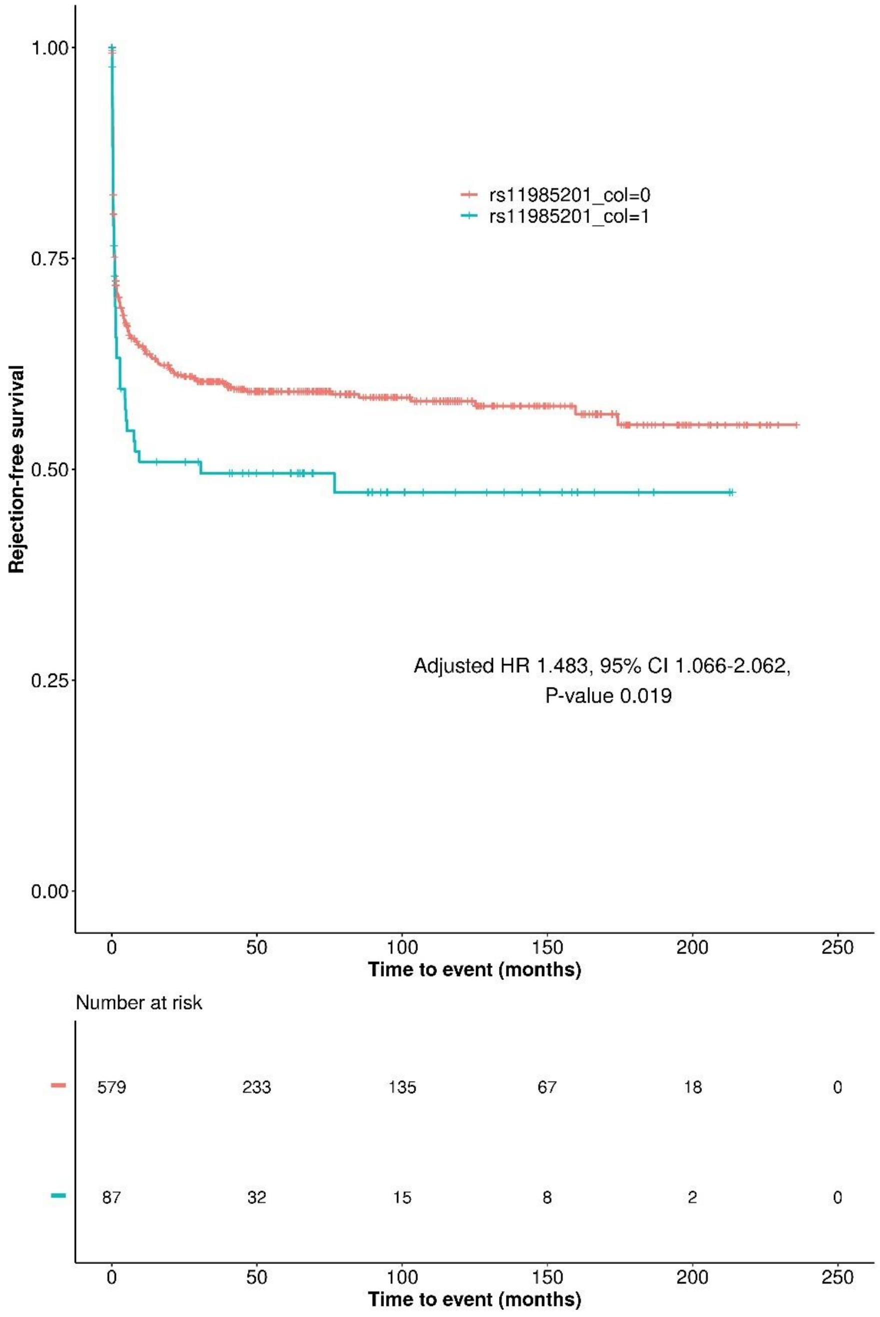

**Supplementary Figure S10. Kaplan-Meier plot displaying the effect of deletion-tagging variant rs11985201 mismatch on acute rejection-free survival.** The 'deletion mismatch' group with recipients who were homozygous for the deletion-tagging allele A and received a transplant from a donor with AG or GG is represented in blue. The 'no mismatch' group is represented in orange. Under the survival curve, a risk table is displayed with the number of recipients at risk of acute rejection. The presented hazard ratios, confidence intervals, and *P* values were calculated using the Cox proportional hazards model. The model was adjusted for recipient and donor age, recipient and donor sex, cold ischemia time, HLA I eplet mismatch, HLA II eplet mismatch, year of transplantation, autoimmune liver disease, and initial calcineurin inhibitor type. Affected genes: ADAM metallopeptidase domain 3A (pseudogene) and domain 5 (pseudogene) (*ADAM3A/ADAM5P*). CI, confidence interval; HR, hazard ratio.

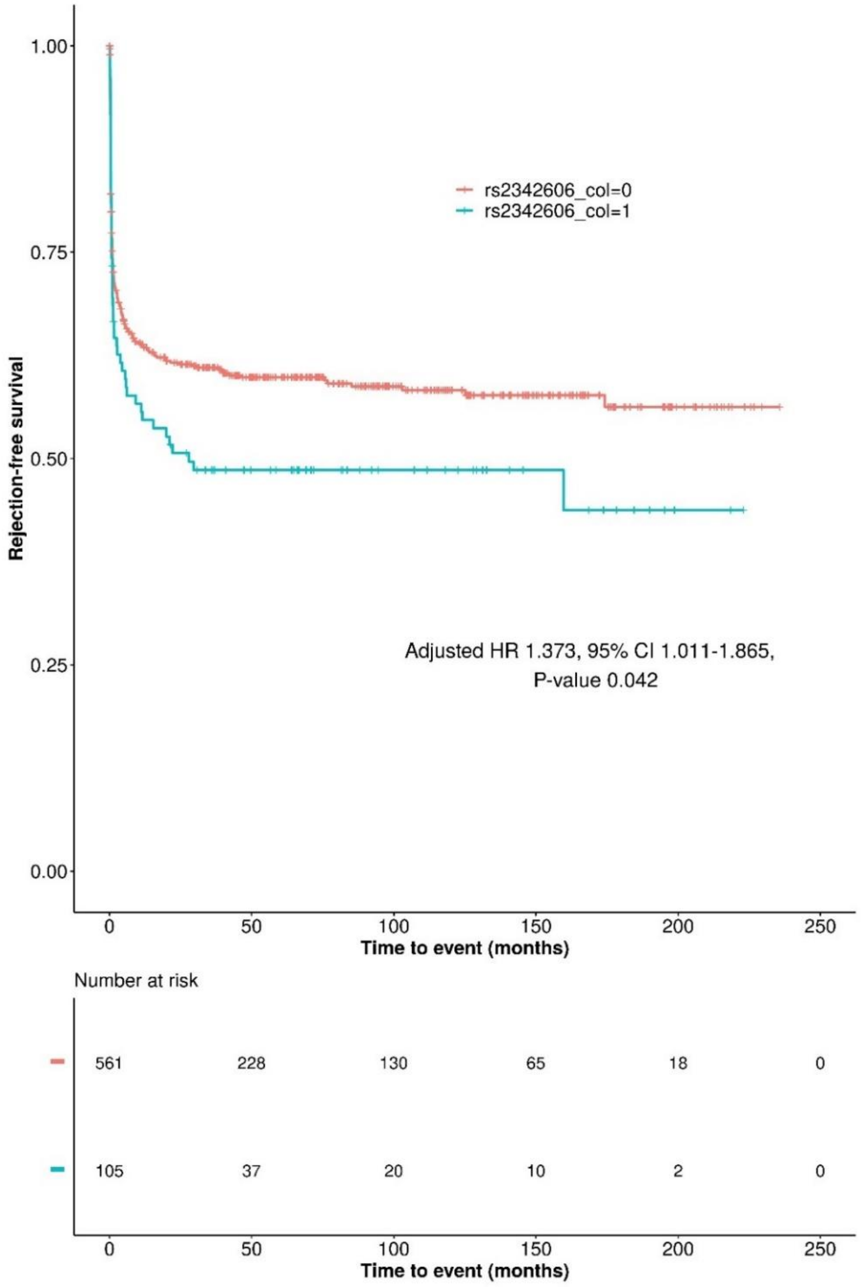

**Supplementary Figure S11. Kaplan-Meier plot displaying the effect of deletion-tagging variant rs2342606 mismatch on acute rejection-free survival.** The 'deletion mismatch' group with recipients who were homozygous for the deletion-tagging allele T and received a transplant from a donor with TC or CC is represented in blue. The 'no mismatch' group is represented in orange. Under the survival curve, a risk table is displayed with the number of recipients at risk of acute rejection. The presented hazard ratios, confidence intervals, and P values were calculated using the Cox proportional hazards model. The model was adjusted for recipient and donor age, recipient and donor sex, cold ischemia time, HLA I eplet mismatch, HLA II eplet mismatch, year of transplantation, autoimmune liver disease, and initial calcineurin inhibitor type. Affected genes: LOC642521 and LOC642538. CI, confidence interval; HR, hazard ratio.

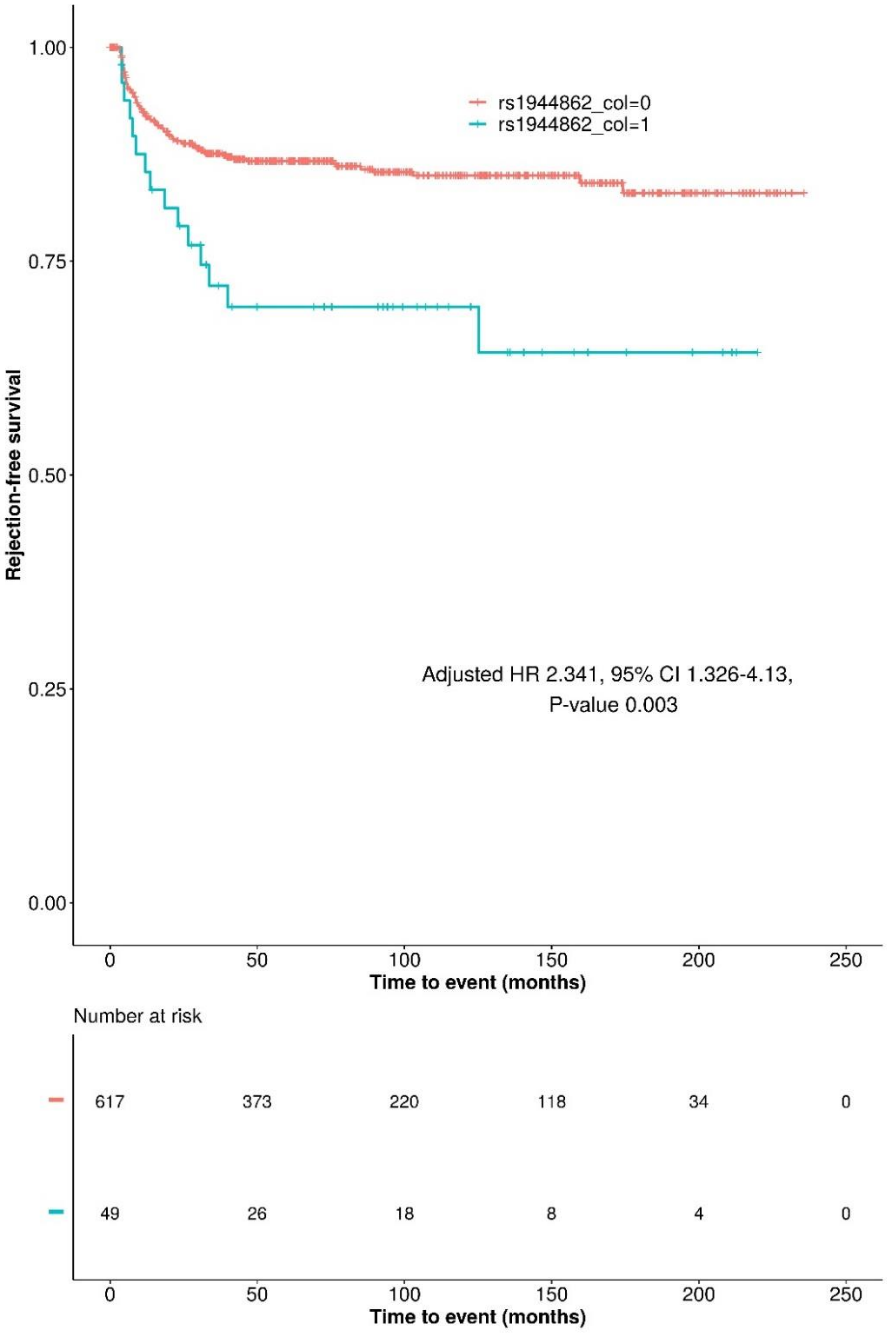

**Supplementary Figure S12. Kaplan-Meier plot displaying the effect of deletion-tagging variant rs1944862 mismatch on late rejection-free survival.** The 'deletion mismatch' group with recipients who were homozygous for a deletion-tagging allele A and received a transplant from a donor with AG or GG is represented in blue. The 'no mismatch' group is represented in orange. Under the survival curve, a risk table is displayed with the number of recipients at risk of late rejection. The hazard ratios, confidence intervals, and *P* values were calculated using the Cox proportional hazards model. The model was adjusted for recipient and donor age, recipient and donor sex, cold ischemia time, HLA I eplet mismatch, HLA II eplet mismatch, year of transplantation, autoimmune liver disease, and initial calcineurin inhibitor type. Affected gene: olfactory receptor family 4 subunit (*OR4P1P*), pseudogene. CI, confidence interval; HR, hazard ratio.

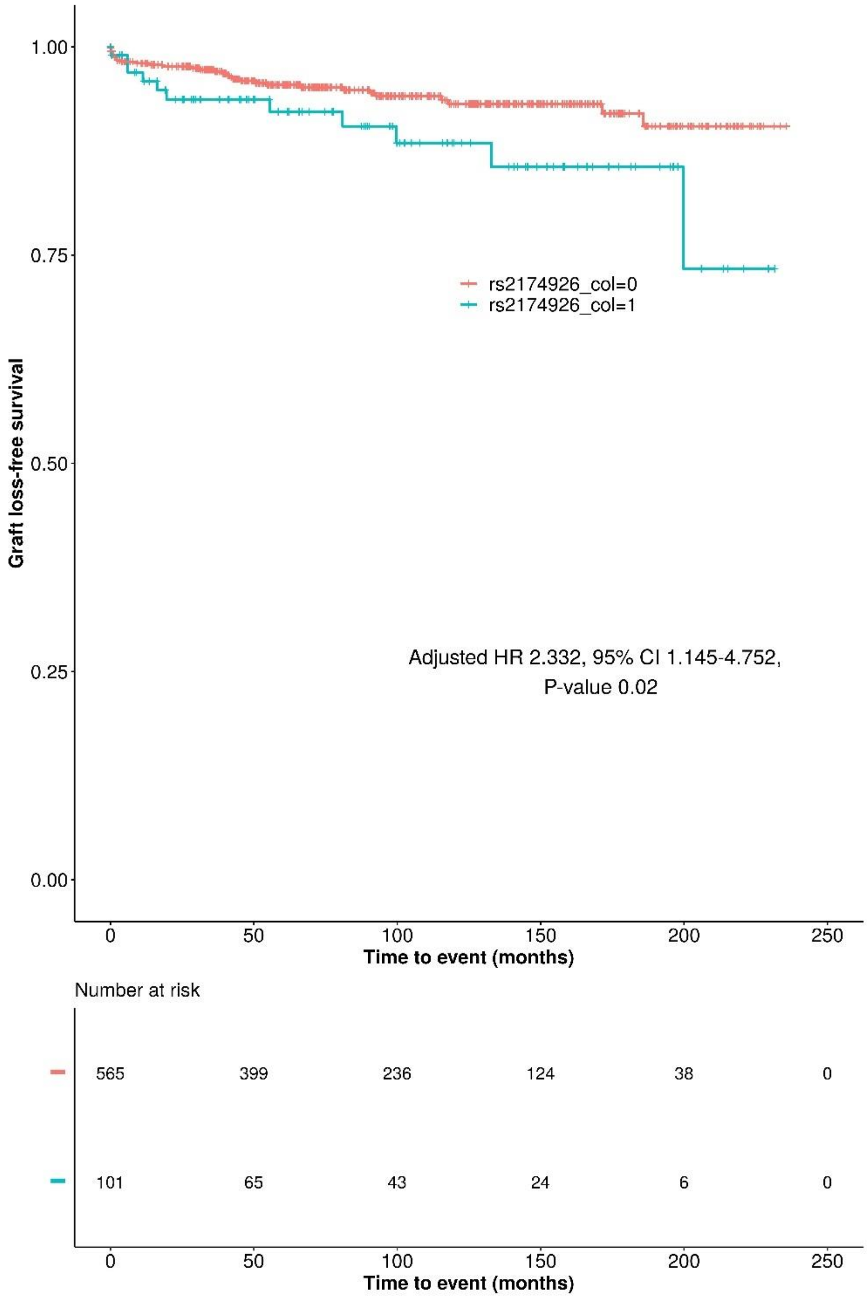

**Supplementary Figure S13. Kaplan-Meier plot displaying the effect of deletion-tagging variant rs2174926 mismatch on graft loss-free survival.** The 'deletion mismatch' group with recipients who were homozygous for a deletion-tagging allele A and received a transplant from a donor with AG or GG is represented in blue**.** The 'no mismatch' group is represented in orange. Under the survival curve, a risk table is displayed with the number of recipients at risk of graft loss. The hazard ratios, confidence intervals, and *P* values were calculated using the Cox proportional hazards model. The model was adjusted for recipient and donor age, recipient and donor sex, cold ischemia time, HLA I eplet mismatch, HLA II eplet mismatch, year of transplantation, autoimmune liver disease, and initial calcineurin inhibitor type. Affected gene: *LOC442434*. CI, confidence interval; HR, hazard ratio.

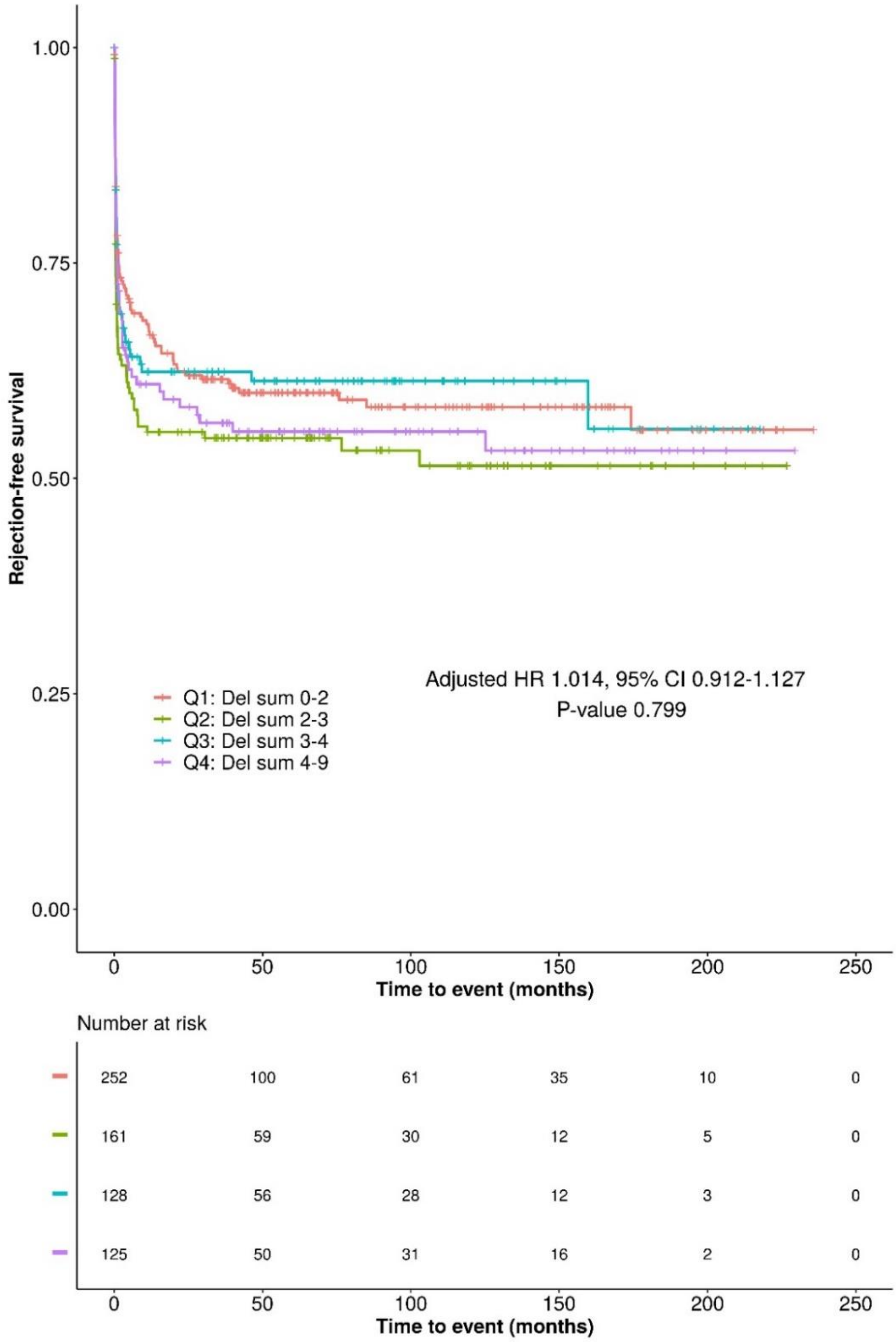

**Supplementary Figure S14. Kaplan-Meier plot displaying the estimated probability of acute rejection -free survival in quartiles of overall deletion mismatch sums between recipients and donors.** Del sum collision value indicates the number of deletion-tagging variant genotype mismatches between recipients and donors. The risk table shows the number of recipients in each quartile group that are at risk of AR at given time points. The presented hazard ratio, confidence intervals and P-value were calculate using Cox proportional hazards model with adjusted covariates: recipient and donor age, recipient and donor sex, cold ischemia time, HLA I eplet mismatch, HLA II eplet mismatch, year of transplantation, autoimmune liver disease and initial calcineurin inhibitor type. Cl, confidence interval; HR, hazard ratio; Q1, quartile 1; Q2, quartile 2; Q3, quartile 3; Q4, quartile 4.

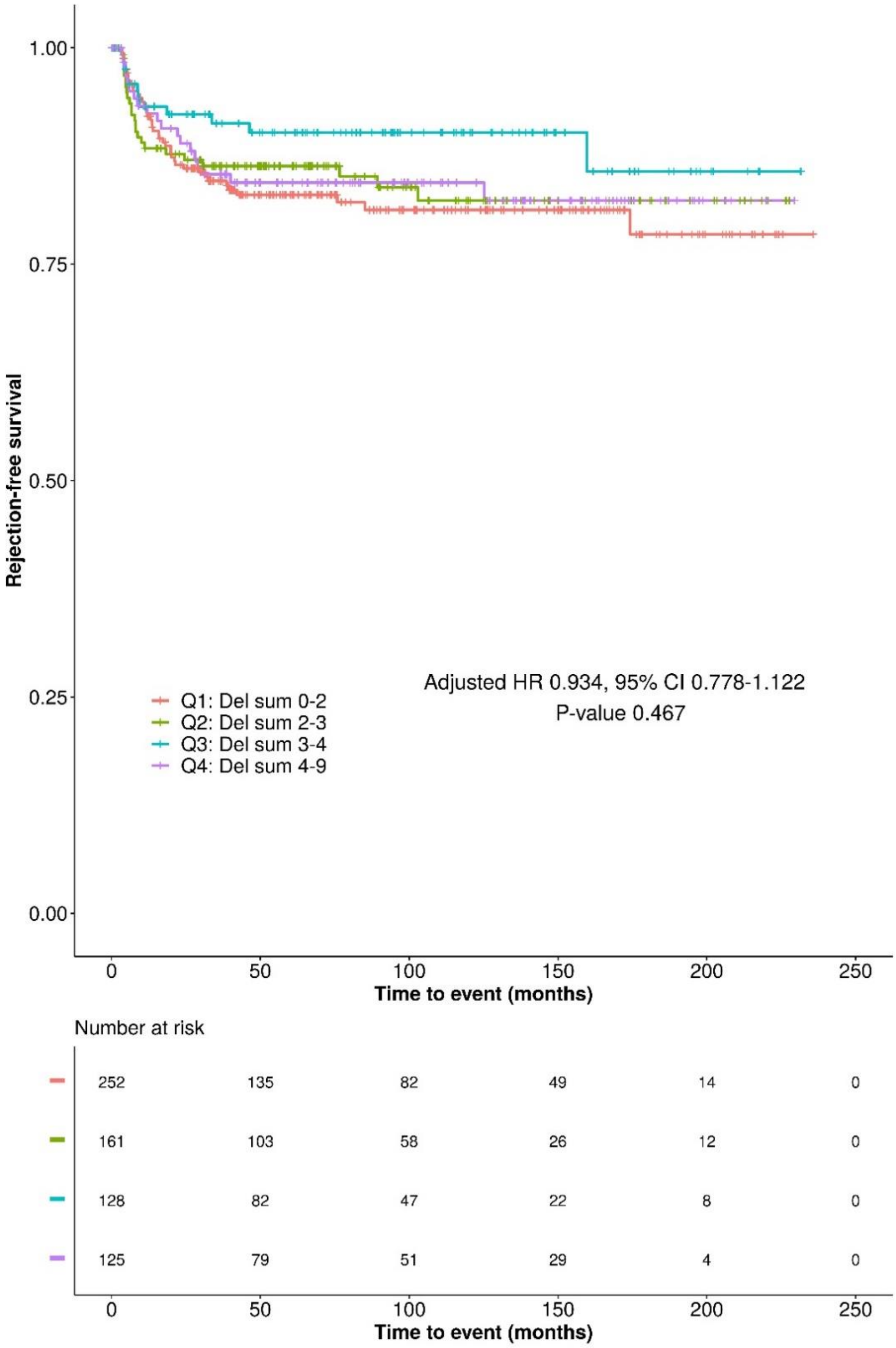

**Supplementary Figure S15. Kaplan-Meier plot displaying the estimated probability of late rejection -free survival in quartiles of overall deletion mismatch sums between recipients and donors.** Del sum collision value indicates the number of deletion-tagging variant genotype mismatches between recipients and donors. The risk table shows the number of recipients in each quartile group that are at risk of late rejection at given time points. The presented hazard ratio, confidence intervals and P-value were calculate using Cox proportional hazards model with adjusted covariates: recipient and donor age, recipient and donor sex, cold ischemia time, HLA I eplet mismatch, HLA II eplet mismatch, year of transplantation, autoimmune liver disease and initial calcineurin inhibitor type. Cl, confidence interval; HR, hazard ratio; Q1, quartile 1; Q2, quartile 2; Q3, quartile 3; Q4, quartile 4.

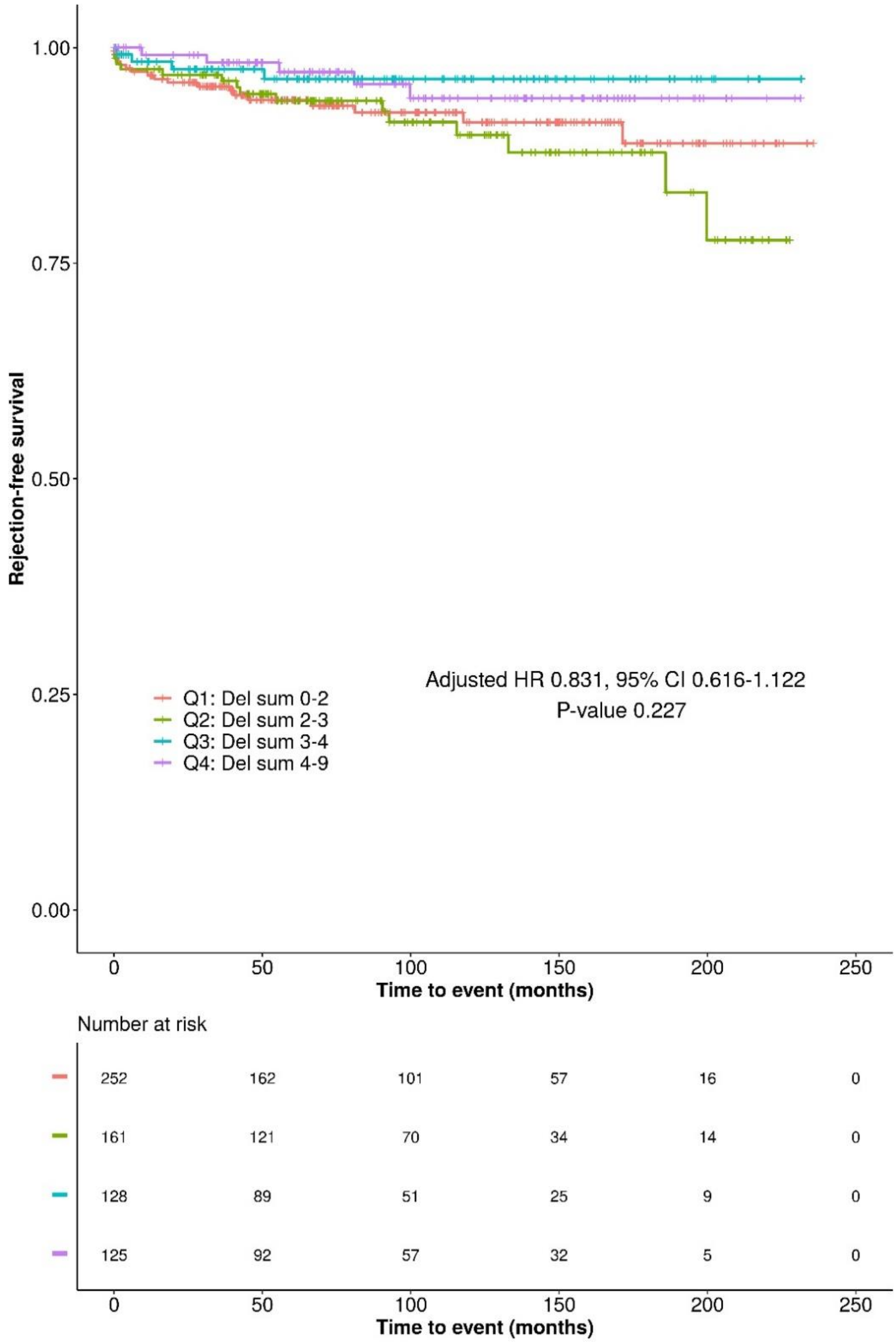

**Supplementary Figure S16. Kaplan-Meier plot displaying the estimated probability of graft loss -free survival in quartiles of overall deletion mismatch sums between recipients and donors.** Del sum collision value indicates the number of deletion-tagging variant genotype mismatches between recipients and donors. The risk table shows the number of recipients in each quartile group that are at risk of graft loss at given time points. The presented hazard ratio, confidence intervals and P-value were calculate using Cox proportional hazards model with adjusted covariates: recipient and donor age, recipient and donor sex, cold ischemia time, HLA I eplet mismatch, HLA II eplet mismatch, year of transplantation, autoimmune liver disease and initial calcineurin inhibitor type. Cl, confidence interval; HR, hazard ratio; Q1, quartile 1; Q2, quartile 2; Q3, quartile 3; Q4, quartile 4.

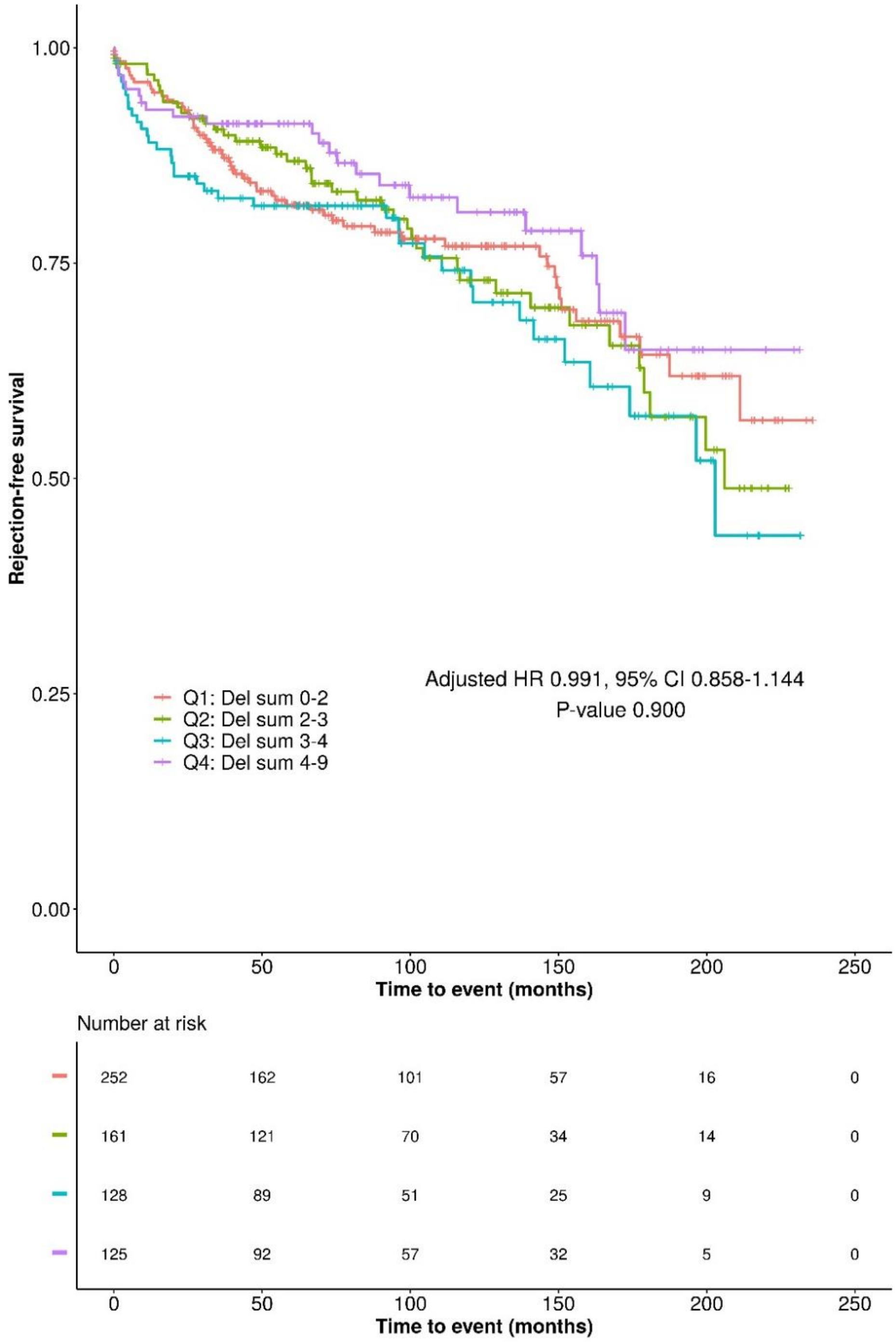

**Supplementary Figure S17. Kaplan-Meier plot displaying the estimated probability of overall survival in quartiles of overall deletion mismatch sums between recipients and donors.** Del sum collision value indicates the number of deletion-tagging variant genotype mismatches between recipients and donors. The risk table shows the number of recipients in each quartile group that are at risk of death at given time points. The presented hazard ratio, confidence intervals and P-value were calculate using Cox proportional hazards model with adjusted covariates: recipient and donor age, recipient and donor sex, cold ischemia time, HLA I eplet mismatch, HLA II eplet mismatch, year of transplantation, autoimmune liver disease and initial calcineurin inhibitor type. Cl, confidence interval; HR, hazard ratio; Q1, quartile 1; Q2, quartile 2; Q3, quartile 3; Q4, quartile 4.
